# Rare schizophrenia risk variant burden is conserved in diverse human populations

**DOI:** 10.1101/2022.01.03.22268662

**Authors:** Dongjing Liu, Dara Meyer, Brian Fennessy, Claudia Feng, Esther Cheng, Jessica S. Johnson, You Jeong Park, Marysia-Kolbe Rieder, Steven Ascolillo, Agathe de Pins, Amanda Dobbyn, Dannielle Lebovitch, Emily Moya, Tan-Hoang Nguyen, Lillian Wilkins, Arsalan Hassan, Psychiatric Genomics Consortium Phase 3 Targeted Sequencing of Schizophrenia Study Team, Schizophrenia Exome Meta-analysis Consortium, Katherine E. Burdick, Joseph D. Buxbaum, Enrico Domenici, Sophia Frangou, Annette M. Hartmann, Dheeraj Malhotra, Carlos N. Pato, Michele T. Pato, Kerry Ressler, Panos Roussos, Dan Rujescu, Celso Arango, Alessandro Bertolino, Giuseppe Blasi, Luisella Bocchio-Chiavetto, Dominique Campion, Vaughan Carr, Janice M. Fullerton, Massimo Gennarelli, Javier González-Peñas, Douglas F. Levinson, Bryan Mowry, Vishwajit L. Nimgaokar, Giulio Pergola, Antonio Rampino, Margarita Rivera-Sanchez, Sibylle G. Schwab, Dieter B. Wildenauer, Mark Daly, Benjamin Neale, Tarjinder Singh, Michael C. O’Donovan, Michael J. Owen, James T. Walters, Muhammad Ayub, Anil K. Malhotra, Todd Lencz, Patrick F. Sullivan, Pamela Sklar, Eli A. Stahl, Laura M. Huckins, Alexander W. Charney

## Abstract

Schizophrenia is a chronic mental illness that is amongst the most debilitating conditions encountered in medical practice. A recent landmark schizophrenia study of the protein-coding regions of the genome identified a causal role for ten genes and a concentration of rare variant signals in evolutionarily constrained genes^1^. This study -- and most other large-scale human genetic studies -- was mainly composed of individuals of European ancestry, and the generalizability of the findings in non-European populations is unclear. To address this gap in knowledge, we designed a custom sequencing panel based on current knowledge of the genetic architecture of schizophrenia and applied it to a new cohort of 22,135 individuals of diverse ancestries. Replicating earlier work, cases carried a significantly higher burden of rare protein-truncating variants among constrained genes (OR=1.48, p-value = 5.4 × 10^−6^). In meta-analyses with existing schizophrenia datasets totaling up to 35,828 cases and 107,877 controls, this excess burden was largely consistent across five continental populations. Two genes (*SRRM2* and *AKAP11*) were newly implicated as schizophrenia risk genes, and one gene (*PCLO*) was identified as a shared risk gene for schizophrenia and autism. Overall, our results lend robust support to the rare allelic spectrum of the genetic architecture of schizophrenia being conserved across diverse human populations.

## Main

Schizophrenia (SCZ) is a severe, chronic psychiatric illness associated with lifelong progression and early mortality ^2-4^. The genetic architecture of SCZ has been deeply characterized over the past fifteen years, with clear genetic contribution from common single-nucleotide polymorphisms (SNPs) ^5^, large copy number variants (CNVs) ^6^, and rare protein-truncating variants (PTVs) ^1,7-14^. Amongst the classes of genetic variation linked to SCZ, rare PTVs provide unique value by linking disease risk to individual genes unambiguously. Most recently, the Schizophrenia Exome Sequencing Meta-Analysis (SCHEMA) consortium increased the sequenced sample size to 24,248 SCZ cases and 97,322 controls, consolidated the enrichment of rare PTVs in SCZ cases across genes under strong evolutionary constraint, and identified ten genes with excess burden of rare disruptive variants in cases compared to controls ^1^. When considered alongside earlier studies, these results suggest that with greater sample sizes additional genes will be found to harbor excess rare PTVs in SCZ. Whole-exome sequencing (WES) and whole-genome sequencing (WGS) allow a hypothesis-free approach to risk gene discovery. However, applying these methods at the scale required to achieve the power necessary to confidently link genes to disease remains cost prohibitive. Targeted sequencing of genes chosen through data-driven algorithms that take into account prior knowledge of the genic PTV burden provided by studies such as SCHEMA is an alternative approach to rapidly achieve the required sample size for novel risk gene discovery.

The majority of large-scale human genetics research initiatives to date have failed to include diverse populations. Over 80% of genome-wide association studies (GWAS) participants are of European ancestry, despite this group comprising less than one-fourth of the total human population ^15,16^. Studies of mental illness have contributed to this disparity with almost exclusive European GWAS cohorts despite roughly equal prevalence of psychiatric disorders worldwide ^17^. The limited evidence from SCZ GWAS and CNV studies of non-European populations suggests broadly shared genetic architecture with that of European populations, but ancestry-specific genetic risk factors are also present ^18-23^. So far, studies of rare PTVs of complex human traits have been largely consistent across ancestries ^24-30^, although no studies have yet shown this for SCZ.

Here, to diversify the population profiled in rare PTV studies of SCZ and achieve the power needed to discover novel risk genes, we designed a custom sequencing panel of 161 putative SCZ risk genes and applied it to case-control cohorts totaling 22,135 individuals from diverse ancestries (40% non-European; Figure 1, Table S1). This study, outlined in Figure 1A and hereafter referred to as the Psychiatric Genomics Consortium Phase 3 Targeted Sequencing of Schizophrenia Study (PGC3SEQ), was limited to cohorts that were not part of earlier SCZ sequencing initiatives such as SCHEMA. A data-driven algorithm was used to construct the sequencing panel that considered all current knowledge of the genetic architecture of SCZ, including a preliminary version of the SCHEMA gene-level burden statistics ^31,32^, with the goal of enriching for genes likely to harbor excess rare PTVs in SCZ but did not reach exome-wide significance in earlier studies due to a lack of power. Specifically, this algorithm was based on gTADA ^33,34^, a Bayesian framework which prioritizes genes by integrating genic rare variant burden statistics (from SCHEMA) with gene membership in gene sets that have implicated in SCZ through a variety of approaches (e.g., GWAS, CNV studies) (Figure 1B, Table S2-S3). The exonic regions of the 161 prioritized genes were sequenced on the Ion Torrent platform and rigorous quality control procedures were carried out subsequently (Supplementary Figure S1-S6). Analyses comparing SCZ to controls were limited to rare PTVs (stop-gain, frameshift indels, or essential splicing donor/acceptor) and deleterious missense (placed into tiers based on the MPC score ^35^ (tier 1: MPC>3; tier 2: MPC 2∼3; non-damaging: MPC<2) variants, and synonymous variants were analyzed as a negative control. To maximize power, PGC3SEQ was meta-analyzed with SCHEMA data (available SCHEMA datasets summarized in Table S4 and Figure S7) and sequencing datasets for bipolar disorder and autism. Two broad types of analysis were performed: (1) global enrichment of all constrained genes on the targeted panel (n=80 genes) to demonstrate the overall role of rare disruptive variants in diverse ancestries and (2) gene-level burden test to identify novel SCZ risk genes.

**Figure 1.**
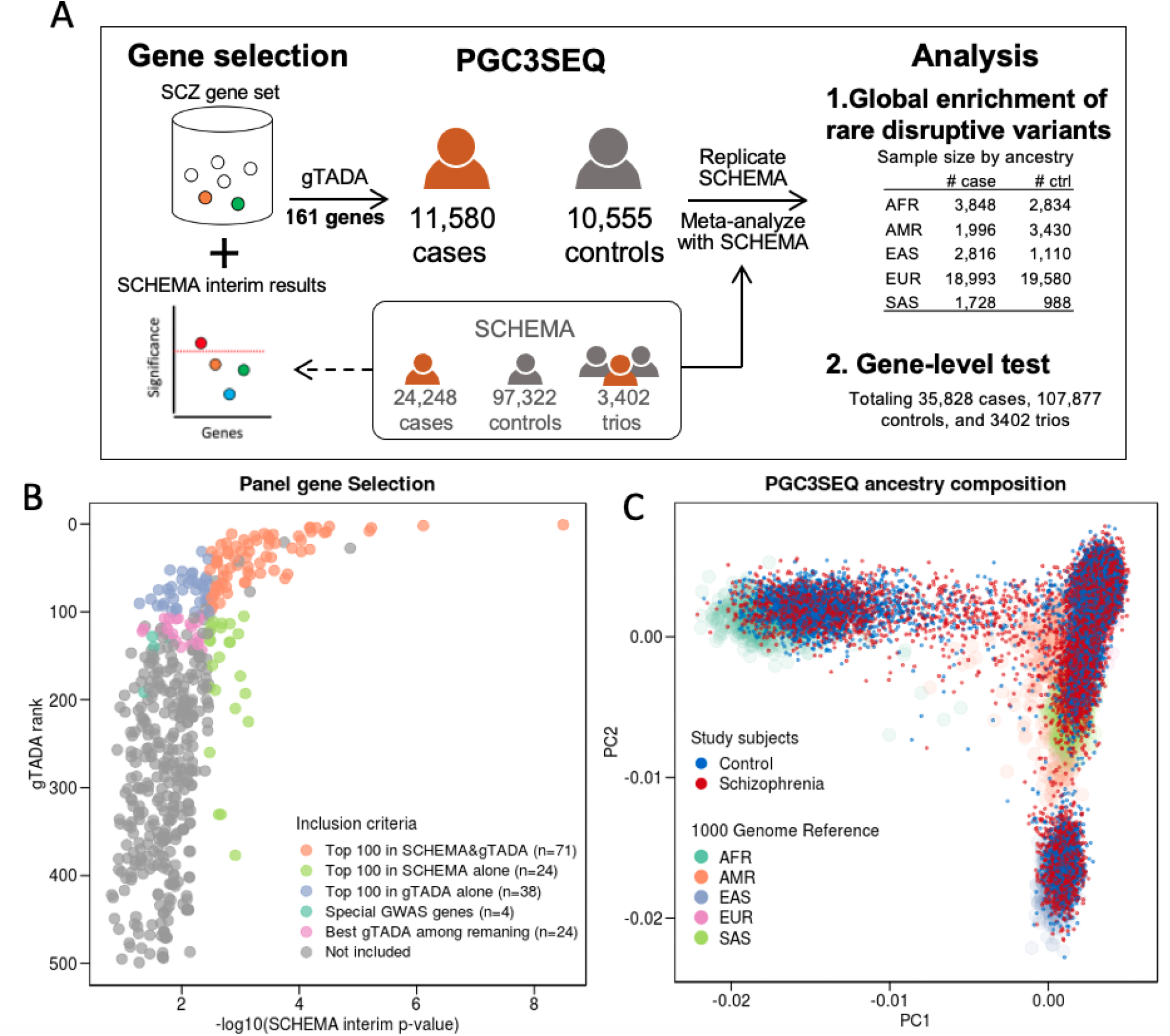
Study design and cohort ancestry composition. (**A**) An overview of the study design. **(B)** Gene selection for the targeted sequencing panel. Genes were selected based on a combination of prior association statistics (SCHEMA), gTADA rankings, and GWAS associations. Specially, we included (1) genes in the top 100 based on the gTADA rank and/or the top 100 based on SCHEMA p-value (Top 100 in SCHEMA&gTADA, Top 100 in SCHEMA alone, Top 100 in gTADA alone, total = 133 genes); (2) genes with evidence for association with SCZ in both GWAS and SCHEMA (special GWAS genes, n=4 genes); (3) an additional 24 genes which had the best 24 gTADA rankings of the remaining genes with a burden p-value < 0.05, to fill up the target panel. X-axis, gene-level p-value using SCHEMA interim data based on which the panel was constructed (different from the final published version); y-axis, gTADA rank of genes. Only the top 500 genes are plotted for a clear display. Some highly ranked genes were excluded (gray dots) due to logistic issues during panel construction. **(C)** PGC3SEQ samples include substantial non-European ancestry. The first two principal components are plotted along the axes, colored by SCZ case control status. 1000 Genome samples are colored by superpopulation. AFR: African, AMR: Admixed American, EAS: East Asian, EUR: European, SAS: South Asian.

PGC3SEQ SCZ cases carried a significantly higher burden of rare PTVs among the 80 constrained genes on the targeted sequencing panel after adjusting for counts of rare synonymous variants and five genotype-derived ancestry PCs (OR=1.48, p-value=5.4 × 10^−6^, Figure 2A, Table S5), replicating in a large independent cohort the excess burden of rare PTVs observed in 3,063 constrained genes in SCHEMA. The higher effect size seen in PGC3SEQ compared to SCHEMA (OR_PGC3SEQ_ =1.48 in 80 genes; OR_SCHEMA_=1.22 in 3063 genes) demonstrates the effectiveness of our gene prioritization strategy. Limiting SCHEMA data to the 80 genes tested in PGC3SEQ showed that the enrichment in PGC3SEQ is much attenuated compared to SCHEMA (OR_PGC3SEQ_=1.48 vs OR_SCHEMA_=3.0, Figure 2A), indicating an effect overestimation in SCHEMA. Tier 1 and tier 2 missense variants were not significantly enriched in SCZ relative to controls in PGC3SEQ. This failure to replicate one of the primary SCHEMA findings may be due to a lack of power, as the effects in two studies were directionally consistent. The burden of rare synonymous variants, which were analyzed as a negative control, was significantly higher in SCZ relative to controls. Sensitivity analysis showed that this signal was due to an overall higher burden of any rare coding variants in SCZ relative to controls, rather than due to technical bias or variability between contributing cohorts (Methods, Figure S8). The global PTV enrichment remained significant after accounting for this overall higher baseline burden (OR=1.4, p-value = 1.2 × 10^−4^, Figure S8C, Table S5).

**Figure 2.**
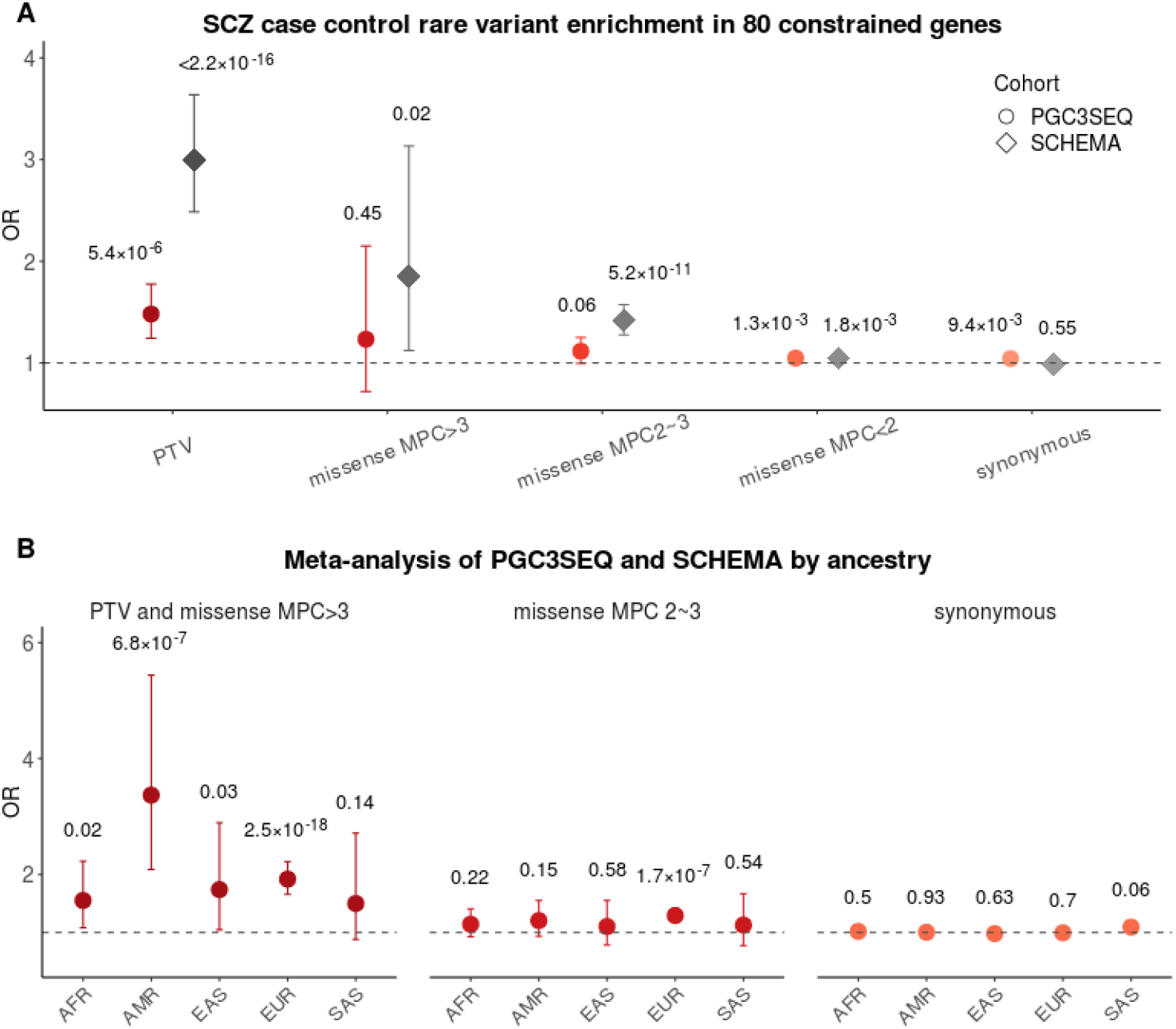
Global enrichment in 80 panel genes under strong constraint (pLI>0.9). (**A**) Case-control enrichment of rare (minor allele count <=5) PTV, missense, and synonymous variants in all ancestries combined, PGC3SEQ results shown in red-orange. We conducted the same analysis in the SCHEMA samples (gray) which we had access to for comparison. The enrichment folds (OR) are on plotted on the y-axis, and bars represent the 95% CIs. P-values were calculated using Firth logistic regression controlling for five ancestry PCs and either rare synonymous variant count (for PTV and missense) or rare non-synonymous variant count (for synonymous variants) to control for potential unknown technical biases. **(B)** Ancestry-stratified rare variant (MAF<0.1%) enrichment in the meta-analysis of PGC3SEQ and SCHEMA. Three groups of variants were analyzed: PTV + MPC>3 missense (combined to increase power); MPC 2∼3 missense; and synonymous variants.

Meta-analysis of PGC3SEQ and SCHEMA were performed to test whether the global enrichment signal was consistent across diverse ancestries (total N=57,323, N by ancestry in Figure 1A). Samples were assigned into five superpopulations used in the 1000 Genome Project (Methods). At the aggregate level, four of the five populations tested displayed a higher burden of rare disruptive variants (PTV + MPC>3 missense) in SCZ cases compared to controls at *p*-value<0.05 (Figure 2B left, Table S6). Although we did not find a nominally significant enrichment in the fifth population (SAS), the magnitude of enrichment was similar to that in the AFR population (OR=1.5), indicating that non-significance is likely due to a lack of statistical power (see power analysis in Methods, Figure S9). When considered separately, PGC3SEQ and SCHEMA provided independent support for the ancestry-stratified enrichments (all ancestries had OR>1 in both datasets, Table S6). Indeed, the PGC3SEQ alone showed nominal significance for AMR, EAS, and EUR, exempt from any potential effect overestimation in SCHEMA. Differences, if any, in the strength of enrichment between pairs of populations were not sizable enough to achieve statistical significance at the current sample sizes. Across five populations, burden of tier 2 missense variants was evaluated although not significant in most (ORs from 1.1 to 1.2, Figure 2B middle), whereas synonymous variants were not enriched in any (Figure 2B right).

Having replicated the global rare PTV enrichment in PGC3SEQ and established its conservation across diverse populations, we then tested for individual genes for harboring excess burden of rare PTVs in SCZ relative to controls. In the PGC3SEQ data alone, none of the 161 genes sequenced were significant after Bonferroni correction (Table S7). The directions of effects of these 161 genes were consistent with the directions observed in SCHEMA (binomial test *p*-value=0.016) and this observation became more pronounced when considering only those 44 genes with a SCHEMA *p*-value < 0.01 (binomial test *p*-value=0.002). Of the ten significant genes identified in SCHEMA, nine were included in the PGC3SEQ panel (*GRIA3* was not). There was an enrichment of rare PTVs on these nine genes collectively (OR=1.66, p=0.03, 49 PTVs in cases vs 24 in controls), and two genes had a p-value<0.05 when considered individually (*RB1CC1* and *CUL1*, Table 1). Notably, *SETD1A*, the gene with the strongest enrichment in SCHEMA, was not replicated in PGC3SEQ, suggesting that its effect size may have been overestimated in SCHEMA (OR_PGC3SEQ_=1.6 vs. OR_SCHEMA_=20.1). Another SCHEMA gene that PGC3SEQ did not support is *CACNA1G*, which among the nine SCHEMA genes on the PGC3SEQ panel had the largest number of PTV events in PGC3SEQ (n=19) yet had an OR of 0.42, directionally inconsistent with its effect in SCHEMA (OR_SCHEMA_=3.1). Despite some evidence of winner’s curse, altogether the gene-level replication tests in PGC3SEQ suggest many of the SCHEMA genes likely confer genuine disease risk, including those not yet reaching the exome-wide level, and their significance might be reached by increasing sample sizes and/or the incorporation of very recent mutations from family data.

**Table 1:** Attempted replication of the nine significant SCHEMA genes in PGC3SEQ

| Gene | PGC3SEQ |  |  |  |  |  | SCHEMA |  |
| --- | --- | --- | --- | --- | --- | --- | --- | --- |
|  | # of PTV allele in case | # of PTV allele in ctrl | # of allele in case | # of allele in ctrl | OR (PTV) | Fisher's exact test p-value | OR (PTV) | P-value |
| *SETD1A* | 9 | 5 | 23160 | 21110 | 1.64 | 0.431 | 20.1 | $2.00 \times 10^{-12}$ |
| *CUL1* | 6 | 0 | 23160 | 21110 | Inf | 0.032 | 36.1 | $2.01 \times 10^{-9}$ |
| *XPO7* | 5 | 0 | 23158 | 21110 | Inf | 0.064 | 52.2 | $7.18 \times 10^{-9}$ |
| *TRIO* | 3 | 3 | 23160 | 21110 | 0.91 | 1.000 | 5.0 | $6.35 \times 10^{-8}$ |
| *CACNA1G* | 6 | 13 | 23160 | 21110 | 0.42 | 0.105 | 3.1 | $4.57 \times 10^{-7}$ |
| *SP4* | 1 | 0 | 23150 | 21104 | Inf | 1.000 | 9.4 | $5.08 \times 10^{-7}$ |
| *GRIN2A* | 0 | 1 | 23152 | 21104 | 0.00 | 0.477 | 18.1 | $7.37 \times 10^{-7}$ |
| *HERC1* | 9 | 2 | 23160 | 21110 | 4.10 | 0.069 | 3.5 | $1.26 \times 10^{-6}$ |
| *RB1CC1* | 10 | 0 | 23148 | 21108 | Inf | 0.002 | 10.0 | $2.00 \times 10^{-6}$ |

Combining SCHEMA and PGC3SEQ (totaling 35,828 cases and 107,877 controls) via a p-value based meta-analysis of gene-level statistics identified two new disease genes at the exome-wide significance threshold (Table 2, Table S7): *SRRM2* (p-value=7.2 × 10^−7^) and *AKAP11* (p-value=4.2 × 10^−7^). In previous work, *SRRM2* has been shown to play a role in the tauopathy of Alzheimer’s disease ^36-38^, and *de novo* mutations in this gene have been linked to developmental disorders ^39^. *AKAP11* was suggested as a trans-gene linking to a SCZ GWAS signal in a recent study ^40^, which considered together with our results, adds to examples of convergence of common and rare variant associations on the same gene. A recent meta-analysis of SCHEMA and a bipolar disorder (BD) dataset also found exome-wide significance for *AKAP11* ^41^, suggesting a role of this gene in the shared etiology of SCZ and BD. The current study consolidates the role of *AKAP11* in SCZ independent of other psychiatric disorders.

**Table 2:** Novel exome-wide significant SCZ genes

| Gene | pLI<sup>a</sup> | PGC3SEQ |  |  | SCHEMA<sup>b</sup> |  | Meta P-value<sup>c</sup> |  |
| --- | --- | --- | --- | --- | --- | --- | --- | --- |
|  |  | # of PTV | OR (PTV) | P-value | OR (PTV) | P-value | SCZ | SCZ & Autism |
| *AKAP11* | 0.98 | 17 | 4.26 | 0.014 | 5.25 | $8.28 \times 10^{-6}$ | $4.15 \times 10^{-7}$ | - |
| *SRRM2* | 1 | 10 | 9.12 | 0.013 | 7.14 | $1.53 \times 10^{-5}$ | $7.19 \times 10^{-7}$ | - |
| *PCLO* | 1 | 8 | 5.01 | 0.024 | 4.02 | $9.36 \times 10^{-4}$ | $1.06 \times 10^{-5}$ | $5.84 \times 10^{-8}$ |

<sup>a</sup> Probability of loss-of-function intolerance
<sup>b</sup> The SCHEMA p-values were retrieved from SCHEMA summary statistics and represent strength of evidence from both case-control and patient-proband trio (*de novo* mutation) data
<sup>c</sup> Meta P-values were determined by Stouffer's method and weighted by sample size. SCZ, meta of PGC3SEQ and SCHEMA; SCZ&Autism, further meta with Autism Sequencing Consortium WES

Lastly, gene-level rare disruptive variant statistics from SCZ, autism spectrum disorder (ASD) ^42^ and BD ^41^ were meta-analyzed to identify pleiotropic risk genes that are not detectable at the sample sizes attained by studies of any single disorder. This identified *PCLO* as a shared risk gene for SCZ and ASD for the first time (*p*-value=5.8 × 10^−8^, Table 2). *PCLO* was not significant in the meta-analysis of SCZ and BD samples; however, this may be due to a lack of power in the BD study (OR_BD_=5.2, p-value=0.12). The association of *PCLO* reported here suggests this gene may be driving the common variant association at nearby loci reported in GWAS of SCZ ^43^ and other psychiatric disorders ^44-47^.

To date, the general lack of case-control exome sequencing studies of non-Europeans has made it difficult to assess the degree to which rare PTV associations are susceptible to the well-known confounding effects of ancestry in GWAS and polygenic prediction studies ^48-52^. Without this knowledge, a complete view of the genetic architecture of complex diseases in human populations cannot be established. Here, we have addressed this gap in knowledge with respect to mental illnesses, showing that in SCZ rare PTV burden is conserved across diverse human populations, and therefore the biological processes disrupted by those genes are likely important in the pathogenesis of SCZ across populations. Larger sample sizes in diverse populations will be vital in further elucidating the rare and common genetic architecture of schizophrenia.

There are limitations to the current study. An interim version of the SCHEMA results was used to construct the targeted sequencing panel, and subsequent changes in the SCHEMA analytical strategy led to differences in gene-level statistics used to build the panel and ultimately in the SCHEMA publication. Specifically, the interim SCHEMA statistics ^31,32^ at the time of panel design did not include *de novo* mutations from trios, used a different strategy to combine PTV and missense variants than that ultimately used in the SCHEMA publication, and were compiled before the incorporation of external Genome Aggregation Database (gnomAD) subjects that doubled the size of controls in SCHEMA (the case cohort in SCHEMA did not change between the interim and final version). Comparing the interim and the published SCHEMA results, gene ranks underwent nontrivial changes, with only 27 overlapping genes between the top 100 lists in the two versions of SCHEMA results. Consequently, our panel likely included more random noise than it would have if panel construction had waited until SCHEMA was complete (64% of panel genes dropped out of top 500 in the published version) and missed some important genes (e.g, *GRIA3*, originally ranked far below 161 yet later changed dramatically to be one of the significant ten). As whole-exome sequencing studies of other diseases approach the sample size achieved for SCHEMA and strategies are considered for how to increase power, the current report offers valuable lessons, and we note that results on datasets as large as 24,000 cases and 50,000 controls can still change substantially as more samples are added. The possibility of such changes makes the targeted panel approach vulnerable, and perhaps WES and WGS as the safest strategies despite their cost.

In summary, rare PTVs have a robust role in SCZ risk, and across ancestries their effect is consistently concentrated in genes under strong evolutionary constraint. The deconvolution of this overall contribution into individual genes, especially those that may display ancestry-specific effects, will require sequencing more individuals of diverse backgrounds. Achieving diversity in human genetic research must be the top priority of the field in order to prevent health disparities from worsening as the findings from genetic research begin to be translated into clinical practice.

## Data Availability

All data produced will be deposited to dbGAP.

