## Supplementary material for "Rare schizophrenia risk variant burden is conserved in diverse human populations": Methods and Supplementary figures

**Methods and supplementary material**

### Methods

#### Cohorts

A brief description of the individual contributing sample collection of PGC3SEQ is available in the supplementary information. To ensure compatibility with Psychiatric Genomics Consortium (PGC) definitions, we define cases as those having a diagnosis of schizophrenia or schizoaffective disorders. A total of 23,352 samples selected to be non-overlapping with SCHEMA as well as other previous and ongoing sequencing efforts in the field were identified and sequenced (Table S1). All participants provided written informed consent before sample collection.

#### Gene panel construction

We intended to build a panel of putative SCZ risk genes from within which the majority of new discoveries from additional WES/WGS would come. To this end, we applied both traditional burden statistics and gTADA to the SCHEMA data. (1) Traditional burden statistics: For each gene in SCHEMA, the enrichment statistics of rare variants in cases compared to controls was calculated using Fisher’s exact test separately for PTVs and damaging missense variants, and then the two classes of variants were combined using meta-analysis to generate a gene-level p-value. Of note, this gene-level p-value is different from that in the SCHEMA publication, which used a slightly different strategy in combining PTVs and missense variants, additionally incorporated evidence from *de novo* mutations using trio data, and included external Genome Aggregation Database (gnomAD) controls. Such analysis strategy changes in the later stage of SCHEMA have led to nontrivial changes in gene rank, which may impact power of our panel to implicate disease genes. (2) *gTADA:* gTADA (Generalized/Gene-set Transmission And De novo Association Test) is a generalized Bayesian framework where *de novo* and rare variant case/control data are integrated with gene-level external information to identify risk genes for neuropsychiatric disorders ^1,2^. We first sought to identify gene sets associated with SCZ in SCHEMA. Through curation of the literature, we identified an initial set of ~160 candidate gene sets. Next, each set was tested independently for association with SCZ in SCHEMA data using gTADA. From all of the sets tested, we identified 27 significantly enriched gene sets. We then calculated a joint enrichment Z-score from the marginal Z-scores and the gene set correlation matrix and kept the 25 gene sets with positive joint Z-scores (Table S2). For each of the 25 sets retained, gene-level statistics (posterior probability of being a risk gene) were then calculated. The genes were then ranked by this metric, and the mean ranking across the 25 ranks was calculated. Combining (1) and (2), genes in the top 100 based on the gTADA mean ranking across the 25 ranks, or top 100 based on the minimum ranking across the 25 ranks, and/or the top 100 based on the burden test were included in the panel (Figure 1B, Table S3, n=139 genes, 6 were later removed due to logistics of designing the sequencing panel). We next included 4 genes with evidence for association with SCZ in both GWAS and SCHEMA, with the criteria being: burden test p-value < 0.05; top 200 rank in gTADA; encompassing index SCZ GWAS index SNP or, if not, located in a GWAS locus with <=10 genes. Finally, an additional 24 genes were chosen for inclusion by taking the best 24 gTADA rankings of the remaining genes with a burden p-value < 0.05.

Based on the observation that gene-level rare single nucleotide variant burden statistics have been consistent across ancestries in a wide range of diseases ^3-8^ ^9^, our targeted panel was expected to have broad utility across ancestries, even though its construction used European-dominant datasets. This is further consolidated by findings from our own ancestry-stratified analysis (Figure 2B).

#### Sequencing and variant calling

Ion AmpliSeq technology is an amplicon-based enrichment method for creating sequencing libraries. We used Ion AmpliSeq Designer version 6.13 to design amplicons that cover the exons of the 161 genes defined based on the Ion hg19 reference. Mean and median percentage of covered base pairs across all exons were 97.7% and 100%. Sequencing of the PGC3SEQ samples was performed on the Ion Torrent platform at Sema4 in between June 2018 and April 2019. Sequencing plates were matched with respect to ancestry and case/control composition whenever possible. The average sequencing depth across all samples was 224x. Sema4 sequencing facility returned to the research team BAM files with flow signal and associated QC metrics. Single sample calling was performed using Torrent Variant Caller (TVC, https://github.com/domibel/IonTorrent-VariantCaller), which is specially optimized to exploit the underlying flow signal information generated by the Ion Torrent sequencing. Sites were left-aligned, normalized and multiallelic sites were split into separate lines using BCFtools v.1.9 (http://samtools.github.io/bcftools/).

**Genotype-level quality control**

We interrogated the call set with respect to a variety of quality control (QC) metrics and implement procedures to ensure rigorous QC standards. In the absence of well-established QC procedures specifically for Ion Torrent data, we drew on the idea of GATK’s Variant Quality Score Recalibration (VQSR) technique and developed a machine learning genotype-level filter based on 177 quality metrics and annotation profiles, including Ion Torrent sequencing metrics such as QUAL, FMT.GQ, FMT.DP, allele-related metrics such as AF, HRUN, MLLD, and coverage and allele frequency from the gnomAD database v2 (https://gnomad.broadinstitute.org). Considering that the majority of SCHEMA data with which we will meta-analyze were generated on the Illumina platform, we calibrated our Ion Torrent targeted sequencing data using a subset of the control samples (N=1347) who had available Illumina WES data. Specifically, we used XGBoost ^10^ to train the classier in 70% of the Ion Torrent-Illumina paired data using Illumina as ground truth. In the remaining 30% test set, the classifier achieved an area under the curve (AUC) = 0.95, an accuracy = 95.3% and a false discovery rate of 4.4% for SNPs, and an accuracy = 99.0% and a false discovery rate of 6.4% for Indels. Applying the trained classifier to the test dataset improved the concordance between Ion Torrent and Illumina calls from 83.1% to 95.7%. We also compared our machine learning classifier with a set of conventional hard filters and confirmed that the classifier performs unanimously better in all metrics considered (sensitivity, specificity, accuracy, false discovery rate).

Applying the machine learning filter to the entire dataset, 83.2% of the calls were retained, and among the passed variants, 96% are SNPs and 4% are Indels. Five out of 919 detected multiallelics passed the filter and were split into multiple biallelic variants. The proportion of calls that passed the filter among samples used for model training and testing (N=1347) and the remaining samples were similar (83.9% vs 83.1% respectively). Likewise, the pass rate among sites that are covered by both Illumina WES capture and our sequencing panel (33.8% of the calls fall into these regions) and sites only covered by our panel were comparable (85.8% vs 81.8%), indicating that the machine learning model generalized well to new samples and new genomic regions.

#### Sample-level and site-level quality control

To identify low-quality and outlier samples, we examined per-sample sequencing quality metrics including number of mapped reads, average read depth across the panel, on-target rate, and uniformity rate. We also examined sample-level call set characteristics, including call rate, inbreeding coefficient, transition-to-transversion ratio at heterozygote sites, heterozygous-to-homozygous call ratio, total number of variants, number of SNPs and Indels, and number of singletons. We visualized the distribution of the above QC metrics (Figure S1) and identified 94 low-quality/outlier samples which met either one of the following criteria: MappedReads<400,000; MeanDepth<40; OnTarget<80; Uniformity<65; MissingCallRate>0.3; Inbreeding_F>0.6; Het_Hom_Ratio<0.6; Total_SNPs<400; Total_Indels<10. All the QC metrics distributed similarly between SCZ cases and controls (Figure S2).

When combining data from single-sample calls, a no-call at a particular site in a particular sample was deemed as homozygous reference genotype if the depth at that site in that sample is greater than 10 and missing otherwise. Lastly, we applied the site-level filters to exclude variants with a missing rate > 10%.

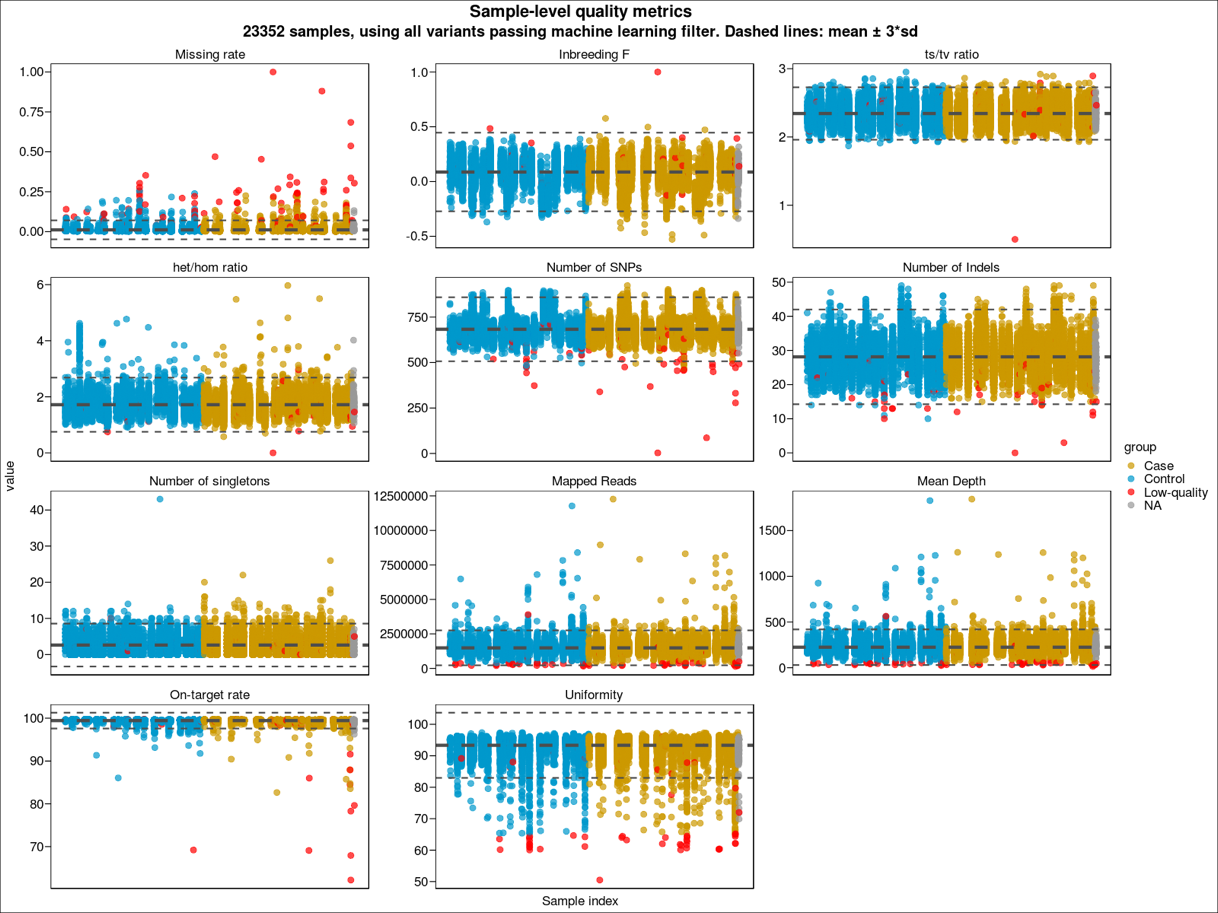

**Figure** **S1**. Sample-level sequencing and calling metrics with outlier and low-quality samples (N=94) highlighted in red.

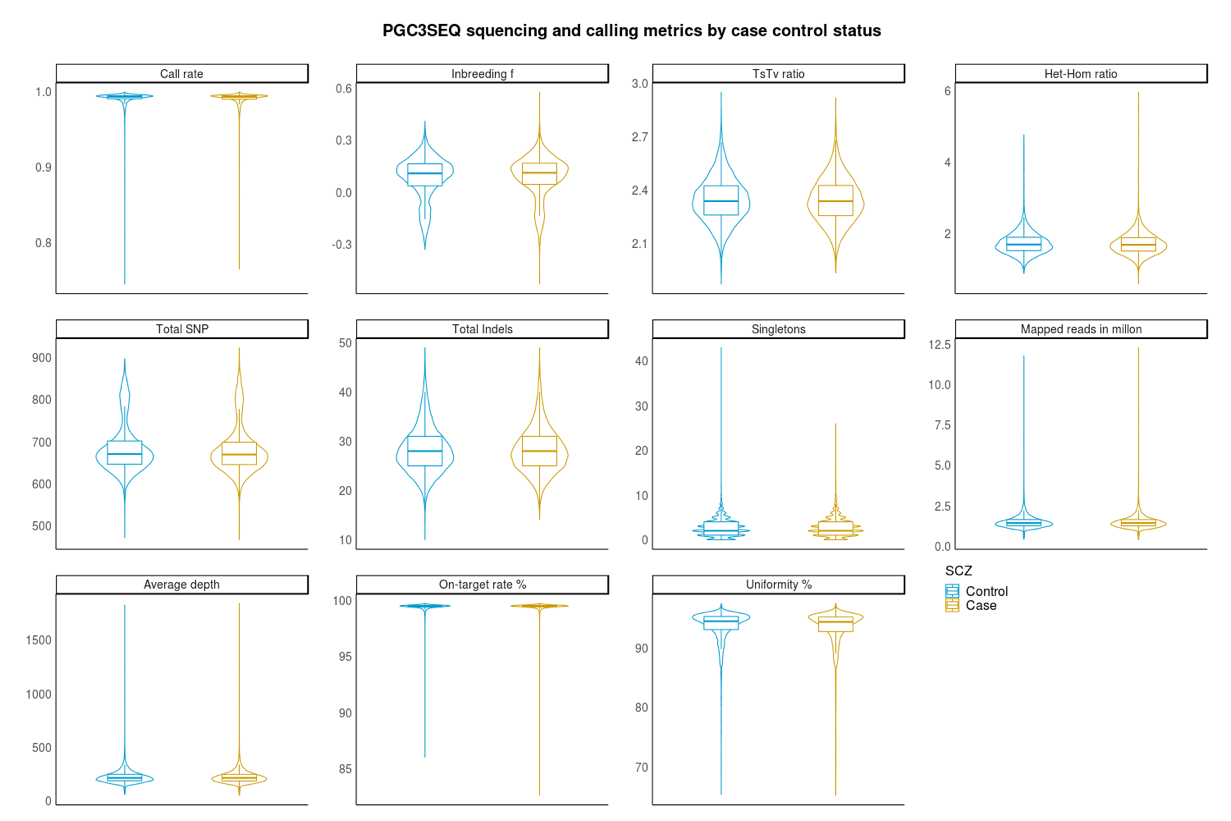

**Figure S2**. Sample-level sequencing and calling metrics by case control status in the final PGC3SEQ sample set.

#### Sample relatedness

We used the population structure adjusted relatedness estimation method PC-AiR and PC-Relate to estimate pairwise relatedness between samples. In addition to the QC steps performed per previous sections, we further LD pruned the dataset and removed Indels before relatedness estimation. Considering that the conventional kinship coefficient ranges for varying degrees of relatedness may not be appropriate when the estimates are from targeted sequencing data covering only a small fraction of the genome, we derived empirical boundaries based on the clustering of sample pairs on an IBS-kinship scatterplot (Figure S3). The unrelated and the related pairs were clearly separated into two clusters with distinct patterns (unrelated pairs: lower oval-shaped cluster, related pairs: upper left). We identified 1096 pairs of genetic relatives and retained one sample from each pairs according to the following prioritization scheme: (1) the sample has fewer genetic relatives in the entire cohort; (2) SCZ patient; (3) the sample has available genome-wide SNP data; (4) the sample has self-reported sex information, (5) the sample has fewer missing genotypes for variants with MAF<0.1%. These measures yielded a total of 22,135 unrelated individuals for downstream analysis.

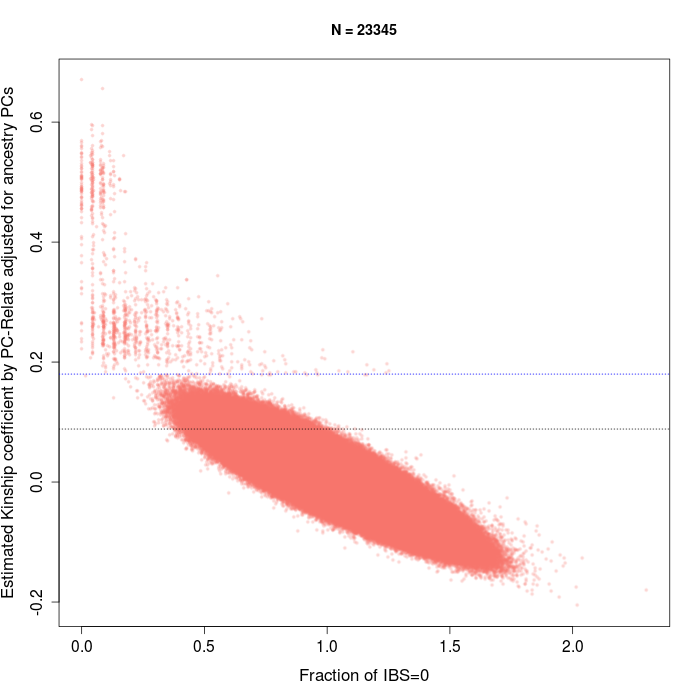

**Figure S3**. Relatedness analysis of 23,352 PGC3SEQ samples using PC-relate. Y axis: kinship coefficients, x axis: the fraction of loci in which individuals share zero alleles IBS. The black (lower) dotted line marks the commonly used lower bound for second-degree relatives (kinship=0.0884). The blue (top) dotted line indicates the customized threshold used for determining unrelated pairs in PGC3SEQ.

#### Description of the final dataset

After all QC steps, the analysis-ready dataset comprised the genetic data of 22,135 unrelated individuals (11,580 cases, 10,555 controls) at 92,813 variants on 161 genes. We compared our call set with two Illumina sequencing datasets (SCHEMA and BioMe in Figure S4), and observed that our dataset is well comparable in terms of SNP/Indels proportions, MAF distribution, and variant annotation type composition.

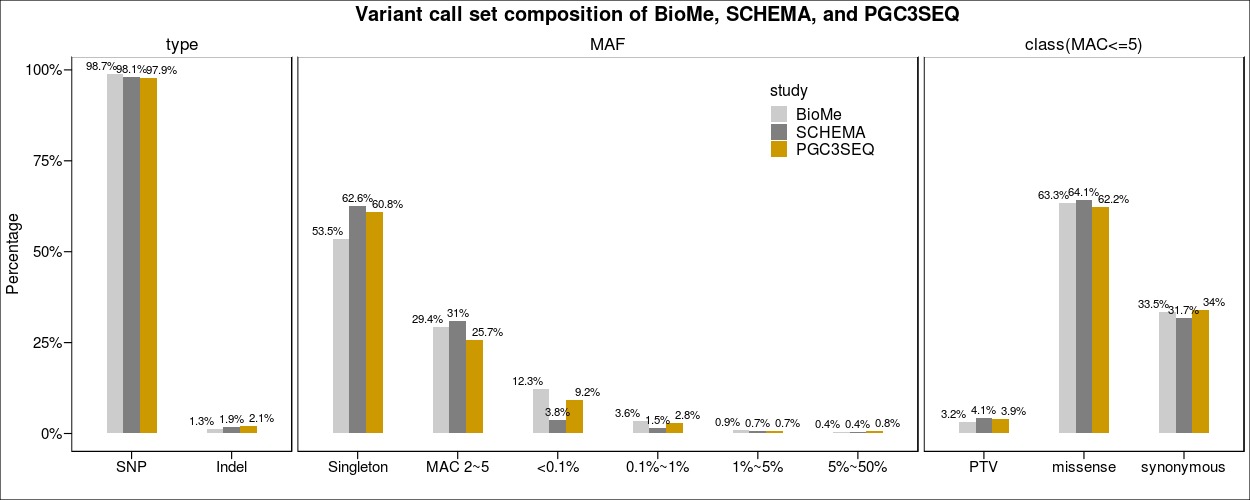

**Figure S4.** Compare call set characteristics of PGC3SEQ with two sequencing datasets generated on the Illumina platform. BioMe consists the whole exome sequencing data of over 30,000 individuals recruited in a health care setting. For BioMe and SCHEMA, we first took the subset of the variants in the 161 genes that are also sequenced in PGC3SEQ. The proportion of SNP/Indel and different MAF bins are calculated in variants of any frequency and any annotation, while the functional annotation comparison is restricted to variants with MAC<=5.

#### Control for population stratification

We calculated ancestry PCs for the 22,135 unrelated individuals in PLINK v1.9 ^11^ using 1,392 LD-pruned common SNPs (MAF>1%) which passed all QC steps. Cases and controls are broadly matched on population structures (Figure S5). The first five PCs were used in later association analysis to control for population substructure.

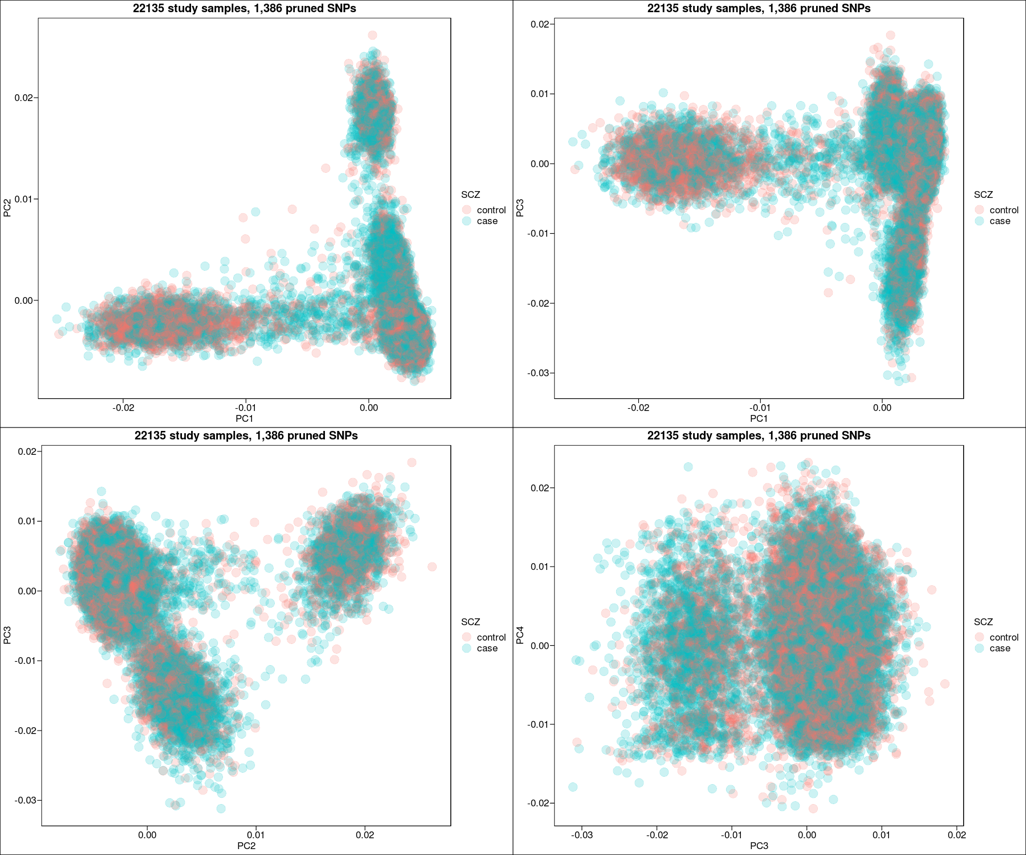

**Figure S5.** First four global PCs for SCZ cases (blue) and controls (red).

#### Ancestry assignment

The genetic ancestry assignment of the PGC3SEQ participants was done by calculating principal components jointly with 1000 Genome phase 3 participants (N=2,501) followed by a K-nearest neighbor classification using the top 3 principal components. We restricted the analysis to 1,372 LD-pruned common SNPs (MAF>1%) that are present in both the study dataset and the reference dataset (1000G). The reference data was first cleaned and quality controlled using PLINK by filtering for missingness per individual (<10%) and missingness per SNP (<10%), and then subsetted to the variant set that passed all the quality control filters in the PGC3SEQ cohort. The cleaned reference and study dataset were harmonized, combined, and pruned for LD, and was then input into PLINK for PCA with default settings.

K-nearest neighbor classification was used for ancestry assignment of the study subjects. Cross-validation determined K=5 and the first three PCs can best classify subjects into five super populations (AFR, AMR, EAS, EUR, SAS). Applying the trained classification model, we assigned each study subject to the super population which includes the most of the subject’s five neighbors. About half our study subjects had self-reported ancestry and ethnicity data, which was broadly consistent with their genetically-inferred ancestry. There is reasonable concordance between country of origin of the sample collection and assigned ancestries (Figure S6).

We then run another round of PCA for each global population separately to generate ancestry-specific PCs, identified ancestry-specific outliers on the PC plots, removed the outliers and re-calculated PCs until no obvious outlier exists. After two rounds of recalculation, 2 EAS and 7 SAS individuals were flagged as outliers within ancestry and were not included in analysis where a stratification by population was performed.

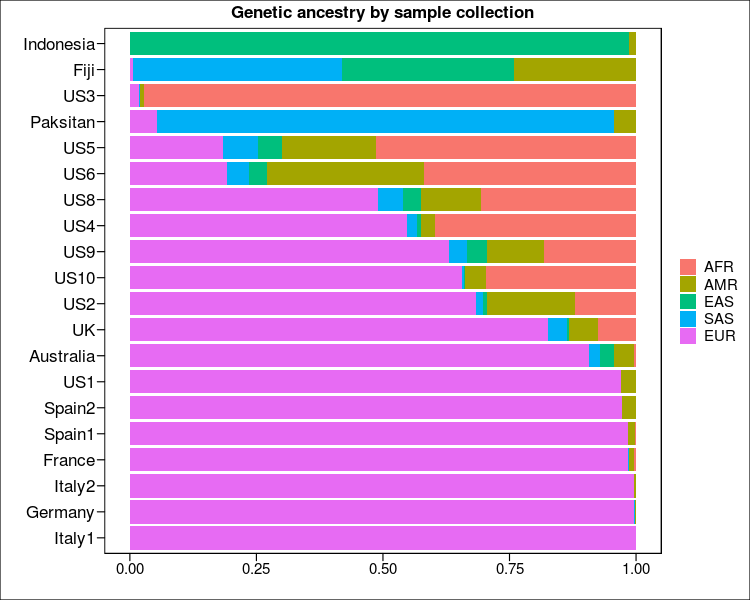

**Figure S6**. Ancestry assignment by individual sample collection. Each sample is assigned a global ancestry by K-nearest neighbor classification using 1000 Genome subjects as a reference. There is reasonable concordance between country of origin and assigned ancestries.

#### Variant annotation

We employed the same variation annotation workflow as used in SCHEMA for the ease of replication and comparison. Specially, annotation by LOFTEE as implemented in the Variant Effect Predictor) ^12^ was applied to variants that passed all QC filters, and the analysis was restricted to the canonical transcript with the most damaging annotation. The three broad types of coding variants analyzed were: (1) protein truncating variants (PTVs), which was defined as any mutation that introduces a stop codon, changes the frame of the open reading frame, or introduces a change at a predicted splice donor or splice acceptor site; (2) missense, which is any single nucleotide variant that causes an amino acid change; or (3) synonymous that results in no amino acid change, as a negative control. Missense variants were further partitioned into groups with increasing deleteriousness based on the MPC (Missense Badness, PolyPhen-2, and Constraint) score annotation ^13^. Tier 1 missense variants have a MPC score > 3, tier 2 has a MPC 2~3, and MPC < 2 indicates non-damaging missense variants. The use of MPC as the missense classifier is based on the SCHEMA results that compared with CADD and PolyPhen, MPC most powerfully prioritized damaging missense *de novo* variants in ASD and DD/ID trios ^14^.

#### Use of SCHEMA data

SCHEMA is a large multi-site collaboration to aggregate, generate, and analyze high-throughput exome sequencing data of SCZ and controls to advance gene discovery. We accessed the post-QC data of a subset of SCHEMA case-control samples with appropriate sharing permissions at the time of this work and did not re-perform genotype-level and sample-level filtering. After excluding 216 samples detected as genetic duplicates with a PGC3SEQ sample, the available SCHEMA datasets contained 19,108 cases and 18,001 controls (Table S4). We used the genetic ancestry label for each individual determined by the SCHEMA analysis team, and within each ancestral group we calculated population-specific principal components using LD-pruned SNPs with MAF>1%, call rate>95, and Hardy-Weinberg p-value>1×10^-6^. Using a similar procedure as used in the PGC3SEQ data analysis, we detected and removed 24 outlier samples from the EAS group. Figure S7 shows the ancestral composition of the SCHEMA cohort, and Table S4 displays the number of SCHEMA cases and controls used for this study by original sample collection.

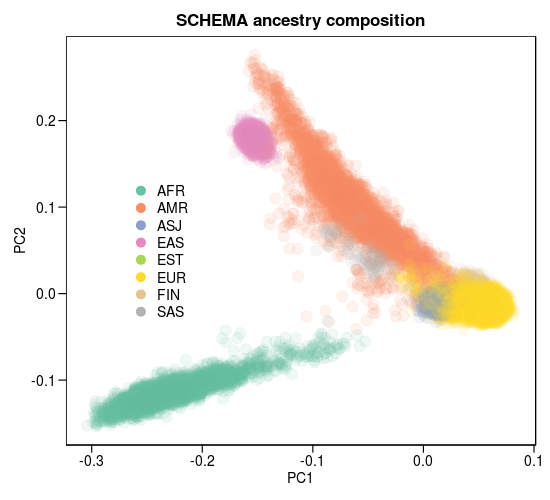

**Figure S7**. Ancestry composition of the SCHEMA samples with available individual-level genetic data. The first two principal components are plotted along the axes, colored by genetically-inferred ancestry. AFR: African, AMR: Admixed American, EAS: East Asian, EUR: European, SAS: South Asian, ASJ: Ashkenazi Jewish, EST: Estonian, FIN: Finnish.

#### Statistical approaches for global enrichment across constrained genes

We defined rare variants as those with minor allele count (MAC) <=5 in the entire sample for any ancestry-combined analysis, and lifted this threshold to MAF<0.1% in ancestry-stratified analysis for a purpose of preserving power. We counted the number of rare variants by annotation type observed in each subject in individual genes and added up the counts across the 80 constrained genes. The association between rare variant burden in the gene set of interest and SCZ status was tested using logistic regression with Firth’s penalized likelihood method to account for sparse data ^15^, while adjusting for ancestry PCs and baseline rare variant burden. The first five global PCs were used in the ancestry-combined analysis, and the first four PCs calculated within each ancestry were used in ancestry-stratified analysis. Baseline rare variant burden was used to control for technical and biological differences between cases and controls. To ensure a minimum correlation between the baseline burden and the burden of interest, we used rare synonymous variant count as the baseline burden when the burden of interest is PTV or missense, and rare non-synonymous variant count as the baseline burden when the burden of interest is synonymous variants. The significance threshold for the enrichment analysis was determined using Bonferroni method correcting for five annotation classes tested (PTV, three missense groups, synonymous), i.e. 0.05/5 = 0.01. A p-value<0.05 was used for nominal significance.

Using the available individual-level SCHEMA data, we performed global enrichment tests across the 80 constrained genes using similar approaches as in the PGC3SEQ analysis. Specifically, we used logistic regression with Firth’s correction and adjusted for ancestry PCs, sex, sequencing cohort, and baseline rare variant burden. The first five global PCs were used in the ancestry-combined analysis, and the first four PCs calculated within each ancestry were used in ancestry-stratified analysis.

Four of the global populations (AFR, AMR, EUR, EAS) had N > 100 in both PGC3SEQ and SCHEMA, and we used inverse-variance weighted meta-analysis to combine their ORs in the two cohorts (sample size by population in Figure 1A). To balance the power reduction due to sample stratification, we relaxed the definition of rare variants to include those with MAF<0.1% (as compared to MAC<=5 in the ancestry-combined analysis). In the full SCHEMA cohort, missense variants with MPC > 3 have a global signal on par with PTVs ^14^, and therefore we grouped these two types of variants together in our analysis of both cohorts to further increase power. Only PGC3SEQ contributed to the analysis of SAS population.

#### Scrutinizing the global enrichment signal of synonymous variants

By definition, synonymous variants and non-damaging missense variants (MPC<2) have no functional consequences on gene products, yet we observed a slightly elevated burden in SCZ cases for these two classes (Figure 2A) which could not be explained by additionally adjusting for sequencing batches or additional lower-rank ancestry PCs. Stratified analysis showed that this signal was not driven by specific sample collections (Figure S8 top) or ancestry groups (Figure S8 bottomleft). When adjusting for the total number of rare coding variants as the baseline burden, the two variant classes were no longer enriched (Figure S8 bottom right), indicating that their signals reflect an overall higher burden of rare coding variants in SCZ cases. This is corroborated by the observation that synonymous variants with a higher allele frequency than those in our main analysis were not enriched (MAF<0.1% synonymous variants had OR=1.00 and p-value=0.98). The PTV signal was robust to the adjustment of the overall rare coding variant burden. The results we reported in the main text for any non-synonymous variants were obtained after adjusting for synonymous variant count, and should be robust to technical or methodological artifacts that would equally affect variants of any annotation.

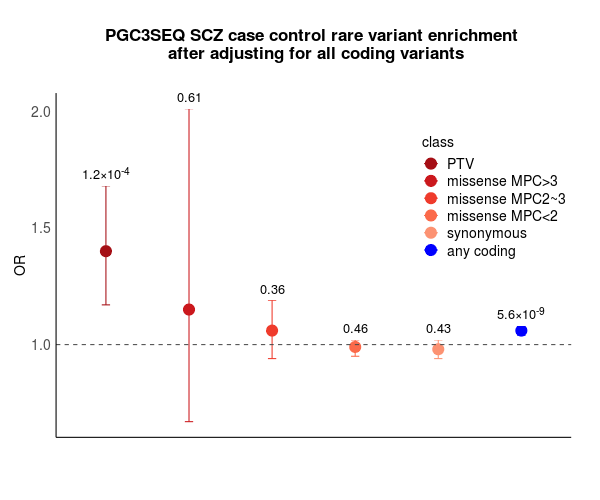

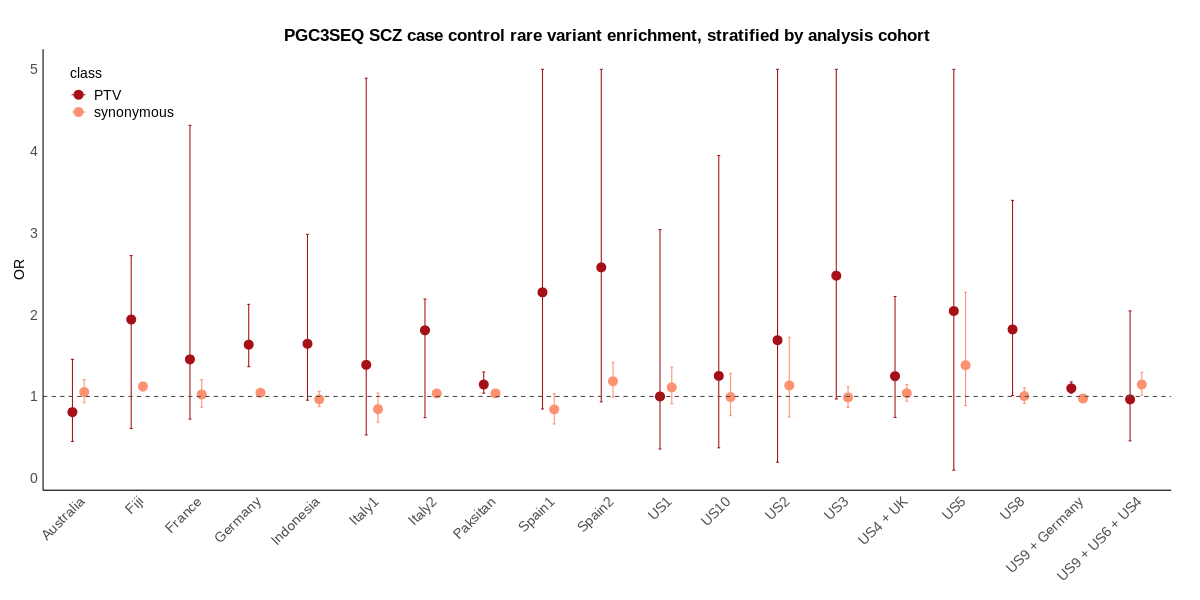

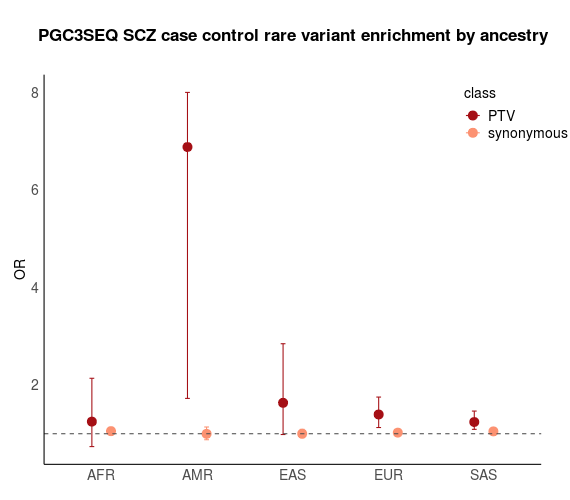

**Figure S8**. Scrutinizing the enrichment signal of synonymous variants. Global enrichment in genes under strong constraint (pLI>0.9) stratified by **(top)** 19 analysis cohorts (some sample collections are combined to form case-control analysis cohorts) and **(bottom left)** five populations. Some sample collections are case-only or control-only and we recombined them to form 19 analysis cohorts. **(Bottom right)** Same as Figure 2A but instead adjusting for the overall burden of rare coding variants across the constrained genes (except for the class of any coding variant which is in blue). The blue dot shows that SCZ cases had a significantly higher overall burden of rare coding variants compared to controls, which gave rise to the enrichment signals of the synonymous and non-damaging missense variants in Figure 2A. After controlling for differences in this background burden, synonymous and non-damaging missense variants no longer showed a signal, while PTVs remained to be enriched at a similar magnitude.

#### Power analysis of the global enrichment test by ancestry

To aid with interpreting our ancestry-stratified analysis results, we conducted Monte Carlo based estimation of statistical power for detecting a significant enrichment signal at different sample sizes, using the osDesign R package ^16^. Figure S9 showed that under a reasonable assumption of a true OR=2, we have adequate power in all population groups except for SAS (power=0.75 in SAS).

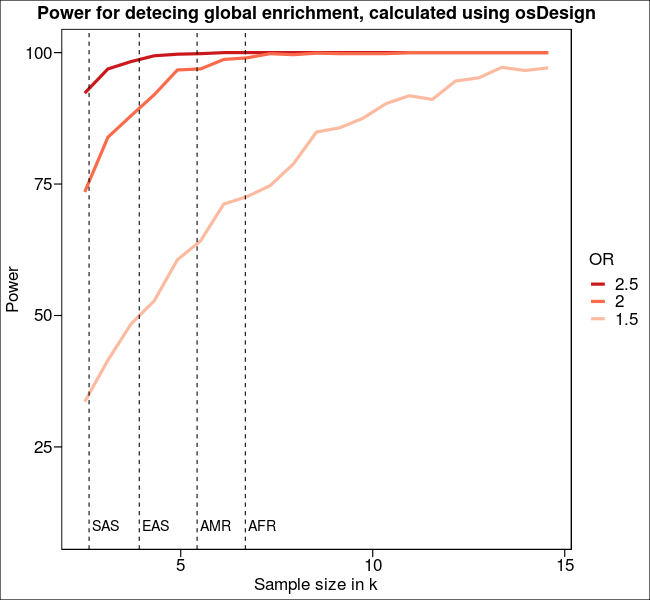

**Figure S9**. Monte Carlo based power estimation of detecting a significant enrichment signal, given three different true ORs. Dotted lines indicate the current sample size for four non-European populations.

#### Statistical approaches for gene-based tests

Gene-based tests aggregate the effects of multiple rare variants and can increase power to detect genetic associations ^17^. It is reasonable to assume that rare disruptive variants in a gene all have the same effect direction (variant alleles associated with higher risk), and under this scenario a burden test is appropriate. Considering the sparsity of the observed count data, we used Fisher’s exact test to compare the burden of PTVs in cases and controls and computed two-sided p-values. The total disruptive burden per gene was quantified by adding up all PTVs (or synonymous variants, as a negative control) annotated to the gene. Different from SCHEMA, we did not incorporate missense variants because they were not significantly enriched globally (Figure 2A). Although Fisher’s exact test is not able to accommodate covariates such as ancestry PCs and baseline burden, this did not adversely affect our analysis as the Q-Q plot showed no sign of inflation in the statistics (Figure S10 top row).

We combined gene-level p-values from PGC3SEQ and SCHEMA (summary statistics obtained from SCHEMA publication) using signed Stouffer’s method, with the sign of the Z scores being the effect direction of the PTVs and weights of each study calculated as:

4 / (1/<# of cases> + 1/<# of controls>) + <# of trios in SCHEMA only>

This meta-analysis totaled 35,828 SCZ cases and 107,877 controls, representing the largest SCZ sequencing dataset to date. The exome-wide significance level was determined as 0.05 / (23,321 tests performed in SCHEMA + 161 tests performed in PGC3SEQ) =2.13 × 10^-6^. As expected, the meta p-values deviate substantially from the null (Figure S10 middle left), consistent with an enrichment of risk genes in the targeted panel. Gene-level synonymous variant p-values displayed the expected null distribution (Figure S10 middle right, Table S8), assuring that the gene-level PTV results were free from technical or methodological artifacts that are agnostic to variant annotation.

We then combined the two SCZ cohorts with the WES datasets of two other psychiatric diseases to identify genes shared across diagnoses. The two studies from which we obtained summary statistics were (1) the latest release of the Autism Sequencing Consortium (ASC) ^18^, and we further converted the gene-level q-values to p-values; (2) the WES of bipolar disorder (BD) by Palmer et al. ^19^. Meta-analysis was performed similarly as above, and the same exome-wide significance threshold was also applied (2.13 × 10^-6^). We noted some degree of control subject overlap between these studies (for example, SCHEMA and ASC both included Swedish control subjects from the same collection). As the overlap between SCHEMA and ASC consist only a small fraction of the entire sample, our analysis (and the discovery of *PCLO*) should only be minimally affected. The controls overlapping between SCZ and BD are expected to be greater per contributing cohort makeup, although we did not identify any new genes.

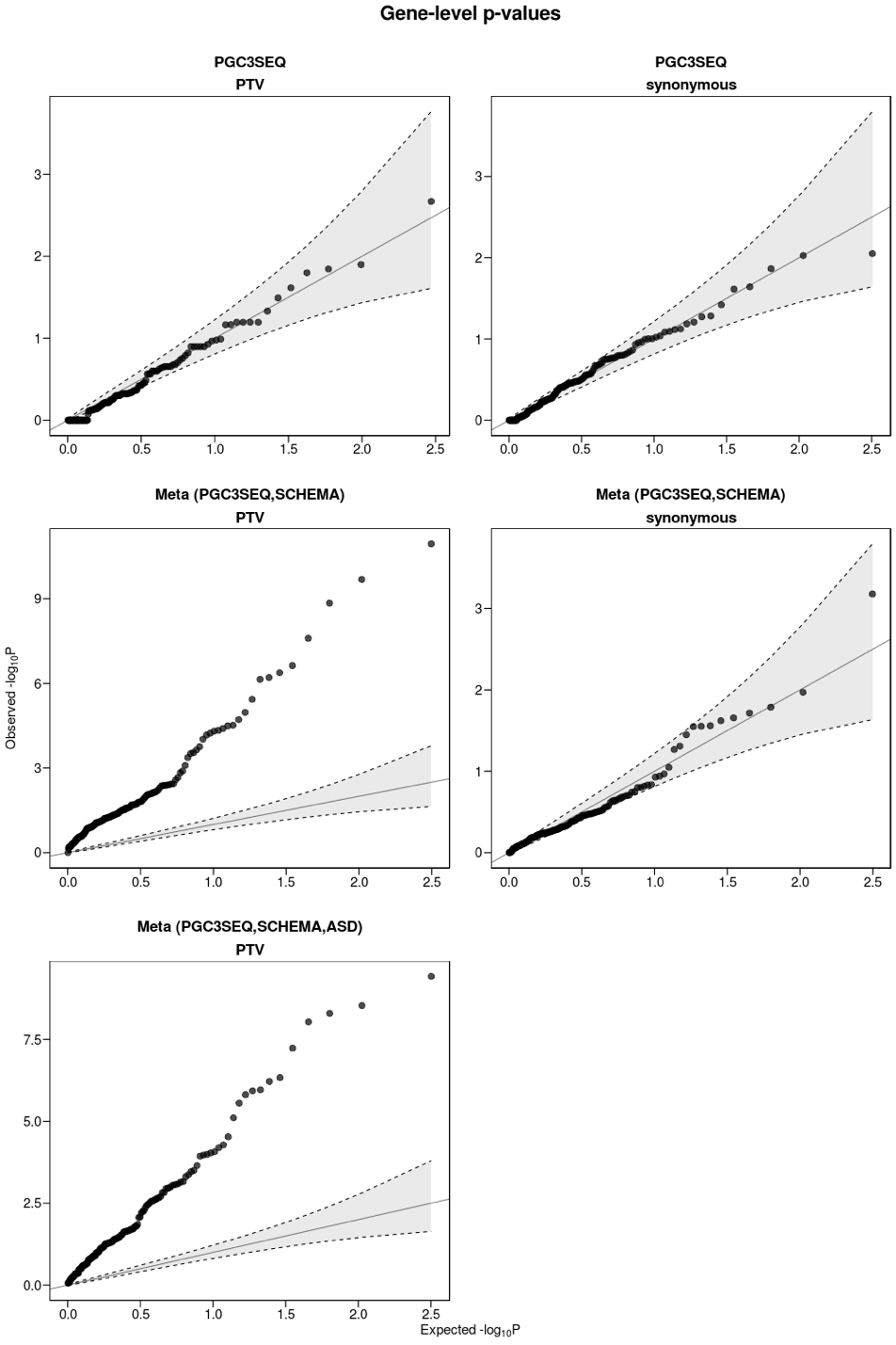

**Figure S10.** *Q*-*Q* plot for gene-based tests of PTV (left) and synonymous variants (right). Observed –log_10_ *P*-values are plotted against expectation given a uniform distribution. Top: PGC3SEQ alone; Middle: Meta-analysis of PGC3SEQ and SCHEMA; Bottom: Meta-analysis of PGC3SEQ, SCHEMA, and the latest WES study of Autism Spectrum Disorder (ASD) ^18^ (summary statistics of synonymous variants were not available for ASD).

### Supplementary information

#### Acknowledgements

A.W.C. is support by the NIMH (R01MH109536)

L.M.H. is supported by the NIMH (R01MH118278; R01MH124839; U01MH109536)

M.C.O., M.J.O., and J.T.R.W. are supported by Medical Research Council Centre Grant No. MR/L010305/1 and Programme MR/P005748/1. For sample acquisition, curation and preparation, we are grateful to Leyden Delta, Magna Laboratories, and their staff (Marinka Helthuis, John Jansen and Adrian King). We also thank Lucinda Hopkins and the core laboratory team at Cardiff University.

K.E.B. is supported by R01MH100125 and 1I01CX000995.

J.D.B. is supported by P50MH066392.

J.M.F. is supported by the Janette Mary O’Neil Research Fellowship, and Australian National Medical and Health Research Council (NHMRC) Project Grant 1063960.

P.R.S. is supported by the Australian National Medical and Health Research Council (NHMRC) Program Grant 1037196 and Investigator Grant 1176716.

D.B.W. and S.G.S. are supported by NHMRC grant 513861.

M.R. has been funded by Instituto de Salud Carlos III projects "PI18/00238" and “PI18/00467” (Co-funded by European Regional Development Fund / European Social Fund "A way to make Europe" / "Investing in your future").

C.A. has received support from the Spanish Ministry of Science and Innovation, Instituto de Salud Carlos III (PI19/024), co-financed by ERDF Funds from the European Commission, “A way of making Europe”, CIBERSAM, Madrid Regional Government (B2017/BMD-3740 AGES-CM-2), and European Union H2020 Program under the Innovative Medicines Initiative 2 Joint Undertaking (grant agreement No 115916, Project PRISM, and grant agreement No 777394, Project AIMS-2-TRIALS), Fundación Familia Alonso, and Fundación Alicia Koplowitz.

M.G. and the work at IRCCS Centro S. Giovanni di Dio, Fatebenefratelli is supported by the Italian Ministry of Health (Ricerca Corrente).

The authors thank the SCHEMA team for sharing their data and results.

#### Completing interests

M.C.O, M.J.O, and J.T.W. are supported by a collaborative research grant from Takeda Pharmaceuticals. A.K.M is a sonsultant at Genomind Inc, InformedDNA. D.M. is a full-time employee of F.Hoffmann-La Roche.C.A. has been a consultant to or has received honoraria or grants from Acadia, Angelini, Biogen, Boehringer, Gedeon Richter, Janssen Cilag, Lundbeck, Medscape, Minerva, Otsuka, Pfizer, Roche, Sage, Servier, Shire, Schering Plough, Sumitomo Dainippon Pharma, Sunovion and Takeda. E.A.S is an employee of Regeneron. The remaining authors declare no competing interests.

#### Detailed cohort description

**US1|Carlos N. Pato, Michele T. Pato**

The cohort recruited participants as part of the Genomic Psychiatry Cohort (GPC), a study based at Rutgers University that recruited controls and cases living and being treated in local communities and healthcare delivery systems. Cases were interviewed using the DI-PAD, a semi-structured clinical interview administered by mental health professionals. Inclusion criteria for cases included meeting lifetime diagnostic criteria for SCZ or schizoaffective disorder in accordance with the OPCRIT algorithms for DSM-IV, ICD-10 or DSM-5 criteria. Individuals reporting no lifetime symptoms indicative of psychosis or mania and who had no first-degree relatives with these symptoms were included as control participants. Exclusion criteria included any premorbid organic mental disorders and premorbid history of significant drug or alcohol dependence by DSM-IV/5 that confounds the diagnosis of SCZ. DNA was extracted from whole blood. All participants gave written informed consent and the IRB of the participating institutions approved the protocol.

**UK|Michael C O'Donovan, Michael J. Owen, James T.R. Walters**

This cohort contains the CLOZUK3 cases who were taking the antipsychotic clozapine and had received a clinical diagnosis of treatment-resistant schizophrenia which in the UK means lack of response to at least two other antipsychotics at standard therapeutic doses for at least 6 weeks. Through collaboration with Leyden Delta, who supply and monitor clozapine in the UK, we acquired DNA from routine blood monitoring samples as previously reported (Pardinas et al, Nature Genetics, 2018). The UK Multicenter Research Ethics Committee approved the study.

**US2|Panos Roussos**

Samples contributed by US2 were derived from three cohorts as part of the CMC study; the Mount Sinai NIH Brain Bank and Tissue Repository, the University of Pennsylvania Brain Bank of Psychiatric illnesses and Alzheimer’s Disease Core Center, and the University of Pittsburgh NIH NeuroBioBank Brain and Tissue Repository. In all cohorts, ethical approval was obtained from all participating sites, and all subjects provided informed consent. Tissue for the collection was dissected at each brain bank and shipped to the Icahn School of Medicine at Mount Sinai (ISMMS) for nucleotide isolation and data generation in one facility to reduce site-specific sources of technical variation. Postmortem tissue from schizophrenia and bipolar disorder cases were included if they met the diagnostic criteria in DSM-IV for schizophrenia or schizoaffective disorder, or for bipolar disorder, as determined in consensus conferences after review of medical records, direct clinical assessments, and interviews of care providers. Cases that had a history Alzheimer’s disease, and/or Parkinson’s disease, or acute neurological insults (anoxia, strokes, and/or traumatic brain injury) immediately prior to death, or were on ventilators near the time of death, were excluded.

MSSM samples - Mount Sinai NIH Brain Bank and Tissue Repository: Brain specimens are obtained from the Pilgrim Psychiatric Center, collaborating nursing homes, Veteran Affairs Medical Centers and the Suffolk County Medical Examiner’s Office. Disease diagnoses are made based on DSM-IV criteria and are obtained through direct assessment of subjects using structured interviews and/or through psychological autopsy by extensive review of medical records and informant and caregiver interviews. Consent is obtained from next of kin. The brain bank procedures are approved by the ISMMS IRB and exempted from further IRB review due to the collection and distributions of postmortem specimens.

Penn samples - University of Pennsylvania Brain Bank of Psychiatric illnesses and Alzheimer’s Disease Core Center: Brain specimens are obtained from the Penn Alzheimer's Disease Core Center prospective collection. Disease diagnoses are made based on DSM-IV criteria and obtained through a clinical interview by psychiatrist and review of medical records. All procedures for Penn are approved by the Committee on Studies Involving Human Beings of the University of Pennsylvania, and the use of control postmortem tissues was considered exempted research in accordance with CFR 46.101 (b), item 65 of Federal regulations and University policy.

Pitt samples - The University of Pittsburgh NIH NeuroBioBank Brain and Tissue Repository: Brain specimens are obtained during routine autopsies conducted at the Allegheny County Office of the Medical Examiner (Pittsburgh) following the consent of the next of kin. An independent committee of experienced research clinicians makes consensus DSM-IV diagnoses for all subjects on the basis of medical records and structured diagnostic interviews conducted with the decedent’s family members. All procedures for Pitt samples have been approved by the University of Pittsburgh’s Committee for the Oversight of Research involving the Dead and Institutional Review Board for Biomedical Research.

**US3| Kerry J. Ressler**

Samples contributed by US3 were part of a larger investigation of genetic and environmental factors in a predominantly African American (AA) urban population of low socioeconomic status with the purpose of determining how those factors may modulate the response to stressful life events. Research participants were approached in the waiting rooms of primary care of a large, public hospital (Grady Memorial Hospital in Atlanta, Georgia) while either waiting for their medical appointments or while waiting with others who were scheduled for medical appointments. Screening interviews, including the participants’ demographic information (e.g., self-identified race, sex, and age) and psychiatric history were completed on site. DNA was extracted from saliva at Mount Sinai. Written and verbal informed consent was obtained for all participants and all procedures in this study were approved by the institutional review boards of Emory University School of Medicine and Grady Memorial Hospital, Atlanta, Georgia.

**Germany| Annette M. Hartmann, Dan Rujescu**

German samples were collected by separate groups within the MooDS Consortium in Mannheim, Bonn, Munich and Jena. In Bonn/Mannheim, cases were ascertained as previously described. Controls were drawn from three population-based epidemiological studies: PopGen, the Cooperative Health Research in the Region of Augsburg (KORA) study, and the Heinz Nixdorf Recall (HNR) study. All participants gave written informed consent and the local ethics committees approved the human subjects protocols. Additional controls were randomly selected from a Munich-based community sample and screened for the presence of anxiety and affective disorders using the Structured Clinical Interview for DSM-IV. Only individuals negative for the above-mentioned disorders were included in the sample.

**US4| Charney W. Alexander**

Founded in September 2007, BioMe is a biobank that links genetic and EMR data for more than 30,000 individuals recruited primarily in ambulatory care settings in the Mount Sinai Health System (MSHS) in New York City. An ethnically diverse, control subset of the BioMe samples was included in the US4 cohort. The current study was approved by the Icahn School of Medicine at Mount Sinai Institutional Review Board (IRB; approval 07-0529). All study participants provided written informed consent.

**US5| Katherine E. Burdick**

US5 consists of adult outpatients with a diagnosis of schizophrenia or schizoaffective disorder who provided informed consent for a cross-sectional study. We collected detailed diagnostic, clinical, and cognitive measures alongside blood collection for DNA and other genetic analyses.

**US6| Sophia Frangou**

Samples contributed by US6 included subjects recruited through the Mount Sinai Conte Center. Ethical approval was obtained from this site, and all subjects provided informed consent.

**US7|Joseph D. Buxbaum**

Samples contributed by US7 were comprised of patients ascertained in Israel. The study was approved by ethics committees in Sheba Medical Center (Israel). All subjects provided informed consent.

**US8|Todd Lencz, Anil K. Malhotra**

Patients with schizophrenia-spectrum disorders (including schizophrenia, schizoaffective disorder, or schizophreniform disorder) were recruited from the inpatient and outpatient clinical services of The Zucker Hillside Hospital, a division of the Northwell Health System. After providing written informed consent, the Structured Clinical Interview for DSM-IV Axis I disorders (SCID, version 2.0) was administered by trained raters. Information obtained from the SCID was supplemented by a review of medical records and interviews with family informants when possible; all diagnostic information was compiled into a narrative case summary and presented to a consensus diagnostic committee, consisting of a minimum of three senior faculty. Healthy comparison subjects were recruited by use of local newspaper advertisements, flyers and community Internet resources, and underwent initial telephone screening to assess eligibility criteria. The nonpatient SCID (SCID-NP) was administered to subjects who met eligibility criteria, to rule out the presence of an Axis I psychiatric disorder; a urine toxicology screen for drug use and an assessment of the subject's family history of psychiatric disorders were also performed. Exclusion criteria included (current or past) schizophrenia spectrum disorder, as well as current Axis I psychiatric disorder, psychotropic drug treatment, or substance abuse. Any subject deemed unable to provide written informed consent was also excluded.

**Pakistan| Muhammad Ayub**

[wait to hear back from contributor]

**US9| Dheeraj Malhotra, Enrico Domenici**

Roche cases were collected by Roche as part of clinical collaborations with hospitals and outpatient centers for eight PhaseII and Phase III, multi-center, randomized, double-blind, parallel-group, placebo-controlled study to evaluate the efficacy and safety of RO4917838 in patients with sub-optimally controlled symptoms of schizophrenia treated with antipsychotics, patients with an acute exacerbation of schizophrenia and in patients with prominent negative and disorganized thought symptoms. Cases were diagnosed according to DSMIV criteria, with medical record review by a trained psychiatrist. Cases gave informed written informed consent, IRBs at each collecting site and Roche ethics committee approved the human subjects protocol.

**Australia| Janice M. Fullerton, Vaughan Carr**

Samples contributed by the ASGC were collected as part of the Australian Schizophrenia Research Bank. Study participants were recruited in four Australian States (New South Wales, Queensland, Western Australia and Victoria) through hospital inpatient units, community mental health services, outpatient clinics and rehabilitation services, non-government mental illness support organizations, and a national multi-media advertising campaign (Loughland et al, ANZJP 2010; PMID:21034186). Ethical approval was obtained from all participating sites, and all participants provided informed consent. A subset of the ASRB sample were employed in a prior paper by the Schizophrenia Working Group of the PGC (Ruderfer et al, Cell 2018; PMID:29906448) under the cohort label "scz_asrb_eur".

**Fiji| Bryan Mowry**

The indigenous Fijian schizophrenia cases and healthy controls included here were part of a wider study to recruit, diagnostically ascertain and collect DNA samples from indigenous Fijian and Fijian Indian participants. Written, informed consent was obtained through procedures approved by the institutional ethics committees and the Fiji Ministry of Health. Cases were recruited from St Giles Hospital, Suva the epicenter for psychiatric services across the Fijian archipelago. Cases included both inpatients and outpatients, who were contacted via an outreach community psychiatric nursing service. Controls were recruited from non-psychiatric outpatient clinics who were screened for mental health issues. 123 indigenous Fijian schizophrenia cases and 26 controls were sequenced for this study.

**France| Douglas F. Levinson, Dominique Campion**

The cohort included two independent French schizophrenia cohorts. Firstly, case participants were unrelated Caucasian in- or out-patients who were recruited for the study of hyperprolinemia in SCZ patients. Cases were interviewed using the PANSS and appropriate sections of the Schedule for Affective Disorders and Schizophrenia, by licensed psychiatrists. Final DSM-III-R diagnoses were assigned by a group of trained psychiatrists based on the interview and available clinical records. We recruited control participants mainly from staff members. The study had ethics approval granted by local IRB ethic committee of Rouen, which permit inclusion of the data in meta-analyses. Blood samples were taken for DNA extraction. A subset of the Rouen- Pitié cohort consisted of 24 cases recruited separately in a study of childhood-onset SCZ in the Paris region. Cases with age at onset between 7-17 years were interviewed using the French version of the DIGS 2.0 by licensed psychiatrists. Participants were included in the genotyping cohort if the final DSM-IV diagnosis was SCZ or schizoaffective disorder depressed type, and the patient and family agreed to provide a blood specimen for genetic studies. The study was approved by the relevant ethics committee.

**Indonesia| Dieter B. Wildenauer, Sibylle G. Schwab**

Samples were obtained through the PsychChip genotyping initiative, and comprised samples from patients with schizophrenia admitted consecutively to psychiatric hospitals in the greater Jakarta area, who were informed about the study and were asked to sign the Informed Consent document for participation in interviews, blood withdrawal and subsequent genetic studies. The study was approved by the institutional review board of the University of Indonesia and by local Ethics Committees. Clinical consensus diagnosis of schizophrenia was made by psychiatrists according to the DSM-IV criteria. Non-psychiatric controls were recruited from students and staff of the University of Indonesia, Jakarta, and from the participating hospitals. Details of the study were previously reported (Schwab et al, Association of rs1344706 in the ZNF804A gene with schizophrenia in a case/control sample from Indonesia. Schiz Res 147: 45-52 (2013)

**Italy1| Massimo Gennarelli, Luisella Bocchio-Chiavetto**

The cohort includes more than 600 schizophrenia patients and not-affected volunteers of Caucasian ancestry for at least two generations, living in the Lombardy region of Italy. The collection of DNA samples and its sharing with the Icahn School of Medicine were approved by the Institutional Ethical Board and authorized by donors under informed consent. Patients admitted to the Psychiatry Unit of Brescia IRCCS Centro S. Giovanni di Dio Fatebenefratelli were enrolled if they had a DSM-IV-TR diagnosis of schizophrenia. Control subjects were randomly recruited from different sources (hospital visitors, cultural and elderly associations, trade unions, etc) and were screened for DSM-IV Axis I disorders. Only volunteers without a history of substance abuse or dependence and without a personal or first-degree family history of psychiatric disorders were enrolled in the study. Subjects who obtained a score lower than 27/30 at the Mini Mental State Examination were excluded as well.

**Italy2| Antonio Rampino, Alessandro Bertolino**

Samples contributed by University of Bari (Bari-Italy) included a total of 1,540 Caucasian white individuals, 913 healthy individuals (487 females) and 627 patients with SCZ (191 females). Ethical approval was obtained from the Local Ethic Committee and all subjects provided informed consent accordingly. Whole blood was collected from all individuals and DNA for subsequent whole genome genotyping was extracted from whole blood.

**Spain1| Celso Arango, Javier González-Peñas**

Samples contributed were derived various cohorts from CIBERSAM (Centro de Investigación Biomédica en Red en Salud Mental, Spain). In all cohorts, informed consent signed by each participating subject or legal guardian and approval from the corresponding Research Ethics Committee were obtained. All of the cohorts, except three of them, were genotyped for a new schizophrenia GWAS at PGC (wave 3).

**Spain2| Margarita Rivera-Sanchez**

Samples contributed by the University of Granada (Spain) were derived from two cohorts. Ethical approval was obtained from both studies, and all participants provided informed consent. Patients were recruited as part of the GENIMS and PISMA studies. GENIMS was a cross-sectional clinical study in which participating patients were consecutive attendees to psychiatric outpatient clinics (Muñoz-Negro J.E. et al, Schizophr Res. 2015; 169(1-3):248-254). All were in a stable stage of their disorder and on antipsychotic medication. Patients were all diagnosed by fully trained psychiatrists using the Structured Clinical Interview for DSM-IV Axis I disorder (SCID-I). Trained raters reviewed these interviews along with available clinical records to determine a consensus lifetime DSM-IV diagnosis of schizophrenia. Additional assessments included sociodemographic and clinical variables such as sex, age, educational level, employment, marital status, and years after onset. To estimate each participant’s premorbid intelligence quotient (IQ), a Spanish version of the Barona index was used. The Spanish version of the PANSS was used to measure psychopathology, since PANSS is the standard scale valid and reliable for this purpose. Global functioning was assessed using the GAF. Inclusion criteria were as follows: 1) meet DSM-IV diagnostic criteria for SZ; 2) be older than 18 years; and 3) agree to participate. Exclusion criteria were as follows: 1) mental retardation and 2) any type of dementia. Control participants were recruited from the PISMA study, which has been reported elsewhere (Cervilla et al., Rev Psiquiatr Salud Ment 2016; 9(4):185-194). This was a cross-sectional study targeting a large representative stratified sample of community-dwelling Andalusian adults between 18 and 75 years of age. All provinces in the Andalusian community were included. Participants were administered the MINI by trained psychologists, which generated both DSM-IV and ICD-10 diagnoses. A saliva sample was obtained from each participant.

**US10| Vishwajit L. Nimgaonkar**

The inclusion criteria for cases were either a diagnosis of schizophrenia or schizoaffective disorder or, at Baltimore, schizophreniform disorder, according to DSM-IV criteria. The cases in Baltimore were recruited from inpatient and day hospital programs of Sheppard Pratt and from affiliated psychiatric rehabilitation programs. The cases in Pittsburgh were recruited from Western Psychiatric Institute and Clinic, Pittsburgh and additional psychiatric treatment facilities in a 500-mile radius of Pittsburgh. Patients were evaluated using structured diagnostic instruments, alongside medical records and informant interviews, where available. The control group was recruited from posted announcements at local health care facilities and universities in the same geographic area and settings where the schizophrenia participants were recruited. All participants provided written informed consent and the study was approved by the Institutional Review Boards of Sheppard Pratt, the University of Pittsburgh School of Medicine, and the Johns Hopkins School of Medicine following established guidelines.

#### PGC3SEQ group author information

Psychiatric Genomics Consortium Phase 3 Targeted Sequencing of Schizophrenia Study Team

Henry S Aghanwa

St Andrew's Toowoomba Hospital, Toowoomba, Queensland 4350, Australia

Moin Ansari

Sir Cowasjee Jehangir Institute of Psychiatry, Hyderabad, Pakistan

Aftab Asif

King Edward Medical University, Lahore, Punjab 54000, Pakistan

Rubina Aslam

Allama Iqbal Medical College, Lahore, Punjab, Pakistan

Jose L Ayuso

Department of Psychiatry, Universidad Autónoma de Madrid, Madrid, Spain

Hospital Universitario de La Princesa, Instituto de Investigación Sanitaria Princesa (IIS Princesa), Madrid, Spain

Centro de Investigación Biomédica en Red de Salud Mental, CIBERSAM, Madrid, Spain

Tim Bigdeli

SUNY Downstate Health Sciences University, Brooklyn, NY 11228, USA

Stefano Bignotti

Psychiatry Unit, IRCCS Istituto Centro S. Giovanni di Dio Fatebenefratelli, 25125 Brescia, Italy

Giuseppe Blasi

University of Bari "Aldo Moro", Bari, Italy

Julio Bobes

Faculty of Medicine and Health Sciences - Psychiatry, Universidad de Oviedo, ISPA, INEUROPA, Oviedo, Spain

Centro de Investigación Biomédica en Red de Salud Mental, CIBERSAM, Madrid, Spain

Bekh Bradley

Department of Psychiatry and Behavioral Sciences, Emory University, Atlanta, GA 30322, USA

Peter Buckley

Virginia Commonwealth University, Richmond, VA 23284, USA

Murray J Cairns

School of Biomedical Sciences and Pharmacy, University of Newcastle, Callaghan NSW 2308, Australia

Hunter Medical Research Institute, Newcastle, New South Wales, Australia

Centre for Brain & Mental Health Research, The University of Newcastle, Callaghan, NSW, Australia

Stanley V Catts

Brain and Mind Centre, The University of Sydney, Sydney, NSW, Australia

School of Medicine, University of Queensland, Herston, QLD Australia

Jorge A Cervilla

Department of Psychiatry, San Cecilio University Hospital, University of Granada, Granada, Spain

Institute of Neurosciences, Biomedical Research Centre (CIBM), University of Granada, Granada, Spain

Abdul Rashid Chaudhry

New Millat Brain Center, Sahiwal District, Punjab, Pakistan

David Cohen

Faculté de Médecine Sorbonne Université, Groupe de Recherche Clinique n°15 - Troubles Psychiatriques et Développement (PSYDEV), Department of Child and Adolescent Psychiatry, Hôpital Universitaire de la Pitié-Salpêtrière, 47-83 Boulevard de l’Hôpital, 75651 Paris Cedex 13, France

Centre de Référence des Maladies Rares à Expression Psychiatrique, Department of Child and Adolescent Psychiatry, AP-HP Sorbonne Université, Hôpital Universitaire de la Pitié-Salpêtrière, 47 - 83 Boulevard de l’Hôpital, 75651 Paris Cedex 13, France

Institut des Systèmes Intelligents et de Robotique (ISIR), CNRS UMR7222, Sorbonne Université, Campus Pierre et Marie Curie, Faculté des Sciences et Ingénierie, Pyramide, Tour 55, Boîte courrier 173, 4 Place Jussieu, 75252 Paris Cedex 05, France

Brett L Collins

Department of Psychiatry, Icahn School of Medicine at Mount Sinai, New York, NY 10029 USA

Angèle Consoli

Faculté de Médecine Sorbonne Université, Groupe de Recherche Clinique n°15 - Troubles Psychiatriques et Développement (PSYDEV), Department of Child and Adolescent Psychiatry, Hôpital Universitaire de la Pitié-Salpêtrière, 47-83 Boulevard de l’Hôpital, 75651 Paris Cedex 13, France

Centre de Référence des Maladies Rares à Expression Psychiatrique, Department of Child and Adolescent Psychiatry, AP-HP Sorbonne Université, Hôpital Universitaire de la Pitié-Salpêtrière, 47 - 83 Boulevard de l’Hôpital, 75651 Paris Cedex 13, France

Javier Costas

Instituto de Investigación Sanitaria (IDIS) de Santiago de Compostela, Complexo Hospitalario Universitario de Santiago de Compostela (CHUS), Servizo Galego de Saúde (SERGAS), Santiago de Compostela, Galicia, Spain

Benedicto Crespo-Facorro

Hospital Universitario Virgen del Rocío, Department of Psychiatry, Universidad de Sevilla, Sevilla, Spain

Centro de Investigación Biomédica en Red de Salud Mental, CIBERSAM, Madrid, Spain

Nikolaos P Daskalakis

Harvard Medical School, Boston, MA 02115, USA

McLean Hospital, Belmont, MA 02478, USA

Michael Davidson

Nicosia University School of Medicine, 2408 Nicosia, Cyprus

Kenneth L Davis

Icahn School of Medicine at Mount Sinai, New York, NY 10029, USA

Faith Dickerson

Sheppard Pratt Hospital, Baltimore, MD 21204, USA

Imtiaz A Dogar

District Headquarter Hospital Failsalbad, Punjab, Pakistan

Elodie Drapeau

Department of Psychiatry, Icahn School of Medicine at Mount Sinai, New York, NY 10029 USA

Lourdes Fañanás

Department of Evolutionary Biology, Ecology and Environmental Sciences, Faculty of Biology, University of Barcelona, Barcelona, Spain

Centro de Investigación Biomédica en Red de Salud Mental, CIBERSAM, Madrid, Spain

Ayman Fanous

University of Arizona, Tuscon, AZ 85721, USA

Veterans Affairs, New York, NY, 10010, USA

Warda Fatima

University of Punjab, Lahore, Punjab, Pakistan

Mar Fatjo

FIDMAG Germanes Hospitalàries Research Foundation, Barcelona, Spain

Departament de Biologia Evolutiva, Ecologia i Ciències Ambientals, Facultat de Biologia, Universitat de Barcelona, Barcelona, Spain

Centro de Investigación Biomédica en Red de Salud Mental, CIBERSAM, Madrid, Spain

Cheryl Filippich

Queensland Brain Institute, The University of Queensland, Brisbane, Queensland 4072, Australia

Queensland Centre for Mental Health Research, University of Queensland, Brisbane, Queensland, Australia

Joseph Friedman

Department of Psychiatry, Icahn School of Medicine at Mount Sinai, New York, NY 10029 USA

John F Fullard

Genetics and Genomics Department, Icahn School of Medicine at Mount Sinai, New York, NY 10029, USA

Penelope Georgakopoulos

Department of Psychiatry and Behavioral Sciences, SUNY Downstate College of Medicine, New York, NY 11203, USA

Marianna Giannitelli

Faculté de Médecine Sorbonne Université, Groupe de Recherche Clinique n°15 - Troubles Psychiatriques et Développement (PSYDEV), Department of Child and Adolescent Psychiatry,Hôpital Universitaire de la Pitié-Salpêtrière, 47-83 Boulevard de l’Hôpital, 75651 Paris Cedex 13, France

Centre de Référence des Maladies Rares à Expression Psychiatrique, Department of Child and Adolescent Psychiatry, AP-HP Sorbonne Université, Hôpital Universitaire de la Pitié-Salpêtrière, 47 - 83 Boulevard de l’Hôpital, 75651 Paris Cedex 13, France

Ina Giegling

Department of Psychiatry and Psychotherapy, Medical University of Vienna, Vienna, Austria

Melissa J Green

School of Psychiatry, University of New South Wales, Sydney, NSW, Australia

Neuroscience Research Australia, Sydney, NSW, Australia

Olivier Guillin

INSERM U1245, 76000 Rouen, Normandie, FranceCentre Hospitalier du Rouvray, Rouen 76000, FranceUFR santé, Université de Rouen Normandie, Rouen, France

Blanca Gutierrez

Department of Psychiatry, Faculty of Medicine, University of Granada, Granada, Spain

Institute of Neurosciences, Biomedical Research Centre (CIBM), University of Granada, Granada, Spain

Herlina Y Handoko

Drug Discovery Group, Cell & Molecular Biology Department, Cancer Programme, QIMR Berghofer Medical Research Institute, Brisbane, Queensland, Australia

Josep M Haro-Abad

Parc Sanitari Sant Joan de Déu, Barcelona, SpainCentro de Investigación Biomédica en Red de Salud Mental, CIBERSAM, Madrid, Spain

Maryam Haroon

Professor Haroon Rashid Clinic, Lahore, Punjab 54660, Pakistan

Vahram Haroutunian

Department of Psychiatry, Icahn School of Medicine at Mount Sinai, New York, NY 10029, USA

Frans A Henskens

School of Medicine and Public Health, University of Newcastle, Newcastle NSW 2308, Australia

Fahad Hussain

Lahore Institute of Research and Development, Lahore, Punjab 54000, Pakistan

Assen V Jablensky

Centre for Clinical Research in Neuropsychiatry, The University of Western Australia, Perth, WA, Australia

Jamil Junejo

Department of Psychiatry and Behavioural Sciences, Liaquat University of Medical and Health Sciences, Jamshoro, Sindh, Pakistan

Brian J Kelly

School of Medicine and Public Health, University of Newcastle, Newcastle NSW 2308, Australia

Dr. Shams-ud-Din A Khan

Al-Shamas Hospital Sargodha, Punjab, Pakistan

Muhammad N S Khan

Queen's University, Kingston, ON K7L 3N6, Canada

Anisuzzaman Khan

Nai Zindage Psychiatric Hospital, Multan, Punjab 60000, Pakistan

Hamid R Khawaja

Azad Jammu and Kashmir Medical College, Muzaffarabad, Azad Jammu and Kashmir 13100, Pakistan

Bakht Khizar

Lahore Institute of Research and Development, Lahore, Punjab 54000, Pakistan

Stella Kim Hansen

Department of Psychiatry and Behavioral Sciences, SUNY Downstate College of Medicine, New York, NY 11203, USA

Steven P Kleopoulos

Genetics and Genomics Department, Icahn School of Medicine at Mount Sinai, New York, NY 10029, USA

James Knowles

SUNY Downstate Health Sciences University, Brooklyn, NY 11228, USA

Bettina Konte

Department of Psychiatry and Psychotherapy, Medical University of Vienna, Vienna, Austria

Agung AAA Kusumawardhani

Department of Psychiatry, cipto mangunkusumo General Hospital, Universitas Indonesia, Jakarta, Indonesia

Claudine Laurent-Levinson

Faculté de Médecine Sorbonne Université, Groupe de Recherche Clinique n°15 – Troubles Psychiatriques et Développement (PSYDEV), Department of Child and Adolescent Psychiatry, Hôpital Universitaire de la Pitié-Salpêtrière, 47-83 Boulevard de l’Hôpital, 75651 Paris Cedex 13, France

Centre de Référence des Maladies Rares à Expression Psychiatrique, Department of Child and Adolescent Psychiatry,AP-HP Sorbonne Université, Hôpital Universitaire de la Pitié-Salpêtrière, 47 - 83 Boulevard de l’Hôpital, 75651 Paris Cedex 13, France

Dannielle Lebovitch

Pamela Sklar Division of Psychiatric Genomics, Genetics and Genomics Department, Icahn School of Medicine at Mount Sinai, New York, NY, 10029, USA

Naeemullah Leghari

Nishtar Medical University, Multan, Punjab 66000, Pakistan

Xudong Liu

Queen's University, Kingston, ON K7L 3N6, Canada

Adriana Lori

Department of Psychiatry and Behavioral Sciences, Emory University, Atlanta, GA 30322, USA

Carmel M Loughland

School of Psychology, University of Newcastle, Newcastle NSW 2308, Australia

Khalid Mahmood

Ar-Rahma Hospital, Multan, Punjab, Pakistan

Saqib Mahmood

University of Health Sciences, Lahore, Punjab 54600, Pakistan

Dolores Malaspina

Icahn School of Medicine at Mount Sinai, New York, NY 10029, USA

Danish Malik

Ameena Clinic, Gujranwala, Punjab, Pakistan

Amy McNaughton

Queen's University, Kingston, ON K7L 3N6, Canada

Patricia T Michie

Vasiliki Michopolous

Department of Psychiatry and Behavioral Sciences, Emory University, Atlanta, GA 30322, USA

Esther Molina

Department of Nursing, Faculty of Health Sciences, University of Granada, Granada, Spain

Institute of Neurosciences, Biomedical Research Centre (CIBM), University of Granada, Granada, Spain

María D Molto

Department of Genetics, University of Valencia, Campus of Burjassot, Valencia, Spain

Centro de Investigación Biomédica en Red de Salud Mental, CIBERSAM, Madrid, Spain

Asim Munir

Wali Neuropsychiatric Center, Faisalabad, Punjab 38001, Pakistan

Gerard Muntané

Hospital Universitari Institut Pere Mata, IISPV, Universitat Rovira i Virgili, Reus, Spain

Centro de Investigación Biomédica en Red de Salud Mental, CIBERSAM, Madrid, Spain

Farooq Naeem

Center for Addiction and Mental Health, Toronto, ON M6J 1H4, Canada

Derek J Nancarrow

Department of Surgery, University of Michigan, Ann Arbor, MI 48109, USA

Amina Nasar

Queen's University, Kingston, ON K7L 3N6, Canada

Tanvir Nasr

Ameena Clinic, Gujranwala, Punjab, Pakistan

Jude U Ohaeri

Department of Psychological Medicine, University of Nigeria Teaching Hospital, Enugu, Enugu State, Nigeria, Africa

Jurg Ott

Laboratory of Statistical Genetics, The Rockefeller University, New York, NY 10065, USA

Christos Pantelis

Melbourne Neuropsychiatry Centre, University of Melbourne & Melbourne Health, Melbourne VIC 3053, Australia

The Florey Institute of Neuroscience and Mental Health, The University of Melbourne, Parkville, VIC, Australia

NorthWestern Mental Health, Melbourne, Vic, Australia

Giulio Pergola

University of Bari "Aldo Moro", Bari, Italy

Sathish Periyasamy

Queensland Brain Institute, The University of Queensland, Brisbane, QLD 4072, Australia

Queensland Centre for Mental Health Research, The University of Queensland, Brisbane, QLD, Australia

Ana G Pinto

BIOARABA Health Research Institute, OSI Araba, University Hospital, University of the Basque Country, Vitoria, Spain

Centro de Investigación Biomédica en Red de Salud Mental, CIBERSAM, Madrid, Spain

Abigail Powers

Department of Psychiatry and Behavioral Sciences, Emory University, Atlanta, GA 30322, USA

Antonio Rampino

University of Bari "Aldo Moro", Bari, Italy

Nusrat H Rana

Punjab Institute of Mental Health, Lahore, Punjab 54000, Pakistan

Mark Rapaport

University of Utah, Salt Lake City, UT, 84112 USA

Abraham Reichenberg

Department of Psychiatry, Icahn School of Medicine at Mount Sinai, New York, NY, USA

James J. Peters VA Medical Center, Bronx, NY 10468, USA

Margarita Rivera

Department of Biochemistry and Molecular Biology II, Faculty of Pharmacy, University of Granada, Granada, Spain

Institute of Neurosciences, Biomedical Research Centre (CIBM), University of Granada, Granada, Spain

Safaa Saker-Delye

Généthon, 1 bis, Rue de l’Internationale, 91000 Evry, France

Ulrich Schall

Priority Centre for Brain & Mental Health Research, The University of Newcastle, Mater Hospital, McAuley Centre, Waratah, New South Wales 2298, Australia

Hunter Medical Research Institute, New Lambton Heights, New South Wales 2305, Australia

Rodney J Scott

School of Biomedical Sciences and Pharmacy, University of Newcastle, Callaghan NSW 2308, Australia

Division of Molecular Medicine, NSW Health Pathology North, Newcastle, NSW 2305, Australia

Megan Shanahan

Department of Psychiatry, Brigham and Women's Hospital, Boston, MA 02115, USA

Cynthia Shannon Weickert

School of Psychiatry, University of New South Wales, Sydney, NSW, Australia

Neuroscience Research Australia, Sydney, NSW, Australia

Department of Neuroscience, SUNY Upstate Medical University, Syracuse, NY 13210, USA

Calvin Sjaarda

Queen's University, Kingston, ON K7L 3N6, Canada

Heather J Smith

Queensland Brain Institute, The University of Queensland, Brisbane, Queensland 4072, Australia

Queensland Centre for Mental Health Research, University of Queensland, Brisbane, Queensland, Australia

Jose Javier Suárez-Rama

Instituto de Investigación Sanitaria (IDIS) de Santiago de Compostela, Complexo Hospitalario Universitario de Santiago de Compostela (CHUS), Servizo Galego de Saúde (SERGAS), Santiago de Compostela, Galicia, Spain

Grupo de Medicina Xenómica, Universidade de Santiago de Compostela, Santiago de Compostela, Spain

Muhammad Tariq

Shafique Psychiatric Clinic, Peshawar, Khyber Pakhtunkhwa, Pakistan

Florence Thibaut

Université de Paris, Faculté de médecine, Hôpital Cochin-Tarnier, Paris 75006, France

INSERM U1266, Institut de psychiatrie et de neurosciences, Paris, France

Paul A Tooney

School of Biomedical Sciences and Pharmacy, University of Newcastle, Callaghan NSW 2308, Australia

Hunter Medical Research Institute, Newcastle, New South Wales, Australia

Centre for Brain & Mental Health Research, The University of Newcastle, Callaghan, NSW, Australia

Muhammad Umar

Lahore Institute of Research and Development, Lahore, Punjab 54000, Pakistan

Elisabet Vilella

Hospital Universitari Institut Pere Mata, IISPV, Universitat Rovira i Virgili, Reus, Spain

Centro de Investigación Biomédica en Red de Salud Mental, CIBERSAM, Madrid, Spain

Mark Weiser

Sheba Medical Center, Tel Hashomer, 52621, Israel

Jin Qin Wu

School of Life and Environmental Sciences, University of Sydney, Sydney, NSW 2006, Australia

Robert Yolken

Stanley Neurovirology Laboratory, Dept of Pediatrics, Johns Hopkins School of Medicine, Baltimore, MD 21205, USA

Ali Zulqarnain

Aleez Neuropsychiatric Centre, Sargodha, Punjab 40100, Pakistan
